# Multi-Criteria Validation of LLM-Inferred Depression Severity from Outpatient Psychiatry Notes

**DOI:** 10.64898/2026.03.11.26348066

**Authors:** Mihael Cudic, William Meyerson, Bo Wang, Qingqing Yin, Pratik N. Khadse, Taylor Burke, Chris J. Kennedy, Jordan W. Smoller

## Abstract

**Background:** Longitudinal measurement of depression severity in outpatient psychiatric care is limited by infrequent standardized assessments. Although psychiatric clinical notes capture illness burden and functional impairment, this information is rarely quantified for analysis.

**Objective:** To evaluate whether large language models (LLMs) can infer clinically meaningful measures of depression severity from outpatient psychiatry notes.

**Methods:** We sampled 91,651 outpatient psychiatry notes from 8,287 adult patients across 58 clinics within a large academic medical center between 2015 and 2021. A HIPAA-compliant LLM (OpenAI GPT-5.2) was prompted to independently estimate three depression severity scores (Patient Health Questionnaire-9 [PHQ-9], Hamilton Depression Rating Scale [HAM-D], and depression-specific Clinical Global Impression-Severity [CGI-S]) from notes, with patient-reported PHQ-9 content within notes redacted to prevent biasing. Convergent validity was assessed against patient-reported PHQ-9 (n=3,757), study-clinician chart review (n=125), and treating-clinician suicide risk assessments (SRA; n=2,985). Predictive validity was evaluated using survival models of antidepressant switching and psychiatric emergency visits. Discriminant validity across diagnoses and consistency across demographic groups and clinics were also evaluated.

**Results:** 10.8% of eligible visits had a PHQ-9 recorded within 7 days before the encounter. LLM-inferred PHQ-9 scores showed moderate agreement with patient-reported PHQ-9 (Cohen’s κ=0.64, 95%CI:0.62-0.66; Pearson r=0.67, 95%CI: 0.65-0.68). Stronger agreement was found between LLM CGI-S and study-clinician chart review (κ_rater1_=0.79, 95%CI: 0.70-0.85; κ_rater2_=0.67, 95%CI: 0.58-0.77; r=0.86 with mean rating, 95%CI: 0.80-0.90). In prospective analyses, LLM CGI-S predicted antidepressant switching (C-index=0.60; CI95%: 0.58-0.62) and psychiatric emergency visits (C-index=0.63; 95%CI: 0.57-0.68), which was comparable to the predictive performance of patient-reported PHQ-9 and treating-clinician SRA. Correlations between LLM CGI-S and patient-reported PHQ-9 were consistent across clinics (I^2^<0.1) but significantly lower among Black (r=0.48, 95%CI: 0.38-0.57) and Hispanic (r=0.43, 95%CI: 0.27-0.56) patients.

**Conclusions:** LLM-inferred depression severity scores from psychiatric outpatient notes support longitudinal, standardized phenotyping of depression severity, such as for routine outcome monitoring. These results have implications for facilitating genetic, pharmacoepidemiologic, and antidepressant treatment effectiveness studies using real-world evidence.

## Introduction

Major depressive disorder (MDD) affects 1 in 5 US adults ^1^ and can markedly reduce quality of life and daily functioning ^2,3^. Electronic health records (EHRs) have become a massive resource for real-world research in depression, supporting applications from genomics to pharmacoepidemiology to treatment effectiveness studies ^4–6^. Many of these applications depend critically on longitudinal measures of symptom severity — for instance, to define treatment response, identify phenotypic subgroups, or adjust for confounding. Yet severity is inconsistently captured in structured EHR data: standardized measures such as the 9-item Patient Health Questionnaire (PHQ-9) appear in fewer than half of psychiatric outpatient visits across healthcare systems ^7^. As a result, EHR-based depression research often relies on indirect proxies — such as changes in billing codes ^8^ or medication regimens ^9^ — that capture clinical decision-making but do not directly reliably measure symptom severity.

Severity measures inferred from narrative clinical notes may provide a complementary source of information as encounter notes are a required element of clinical encounters and describe symptoms, functioning, safety, and any changes since prior visits. Evidence supports high concordance between information in notes and structured reference standards: clinician chart review has shown strong agreement with the Structured Clinical Interview for DSM Disorders (SCID) for some psychiatric diagnoses ^10^. Prior attempts to automatically capture this signal from notes via traditional natural language processing (NLP) have focused on extracting only the presence and absence of MDD ^11^ or written PHQ-9 scores from text ^12^, as inference of severity is highly context-dependent. Moreover, traditional NLP pipelines are labor-intensive, typically requiring hand-crafted feature engineering and extensive manual chart review to generate and validate training labels ^13^.

Large language models (LLMs) may be better suited to depression severity estimation as they represent a major advance beyond traditional NLP in inferring meaning from text in context and, in many domains, can perform at expert level on language understanding tasks ^14–16^. Once validated, LLMs can potentially be applied to new datasets without the hand-crafted pipelines and manual chart review required by traditional approaches ^17^. However, whether current LLM-inferred depression severity scoring approaches are valid enough to support common research or clinical use remains uncertain. The limited evidence available sends conflicting signals: one study found only modest concordance between LLM-inferred and patient-reported PHQ-9 scores (r=0.51; n=15,000) ^18^, while another reported high agreement (κ=0.85; n=77) between LLM estimates and clinician chart review on a Clinical Global Impression Severity scale ^19^. This discrepancy may reflect differences in targets, institutions, or task specification (including which notes are scored); moreover, chart-review agreement alone may not establish validity, because reviewers and models could partly converge on shared documentation artifacts. More fundamentally, available studies have not provided the breadth of validity evidence—convergent, discriminant, and predictive—typically expected before adopting a new severity measure.

We address this gap by evaluating LLM-inferred depression severity from clinical notes across a systematic set of validity tests in a large psychiatric outpatient cohort. We assess convergent validity against multiple independent criteria, including patient-reported PHQ-9, study-clinician chart review, and structured treating-clinician assessments; diagnostic specificity across psychiatric conditions; and predictive validity for subsequent antidepressant changes and psychiatric emergency visits. Together, these analyses provide a comprehensive validation of LLM-inferred depression severity as a measure suitable for EHR-based research. They assess whether routine clinical documentation can support longitudinal severity tracking by extending measurement to visits where standardized patient-reported scales are not collected. Secondarily, we test whether note-based LLM severity—drawing on clinician-documented mental status findings, functional impairment, and clinical assessment—provides complementary prognostic information even when the PHQ-9 is available. Together, these capabilities could enable research applications that require dense longitudinal symptom data currently unavailable in structured EHR fields.

## Methods

### Study Design and Population

We analyzed outpatient psychiatric visit notes from the Massachusetts General Brigham (MGB) healthcare system, drawn from a prior data extraction ^20^ using the Research Patient Data Registry (RPDR). Notes were documented between April 2015 (following system-wide Epic adoption) and November 2021 (end date for prior extraction). Notes from the same provider for the same patient on the same day were concatenated. After excluding group therapy notes, notes with redacted content (flagged as sensitive by the institution), notes authored by non-clinicians, and visits involving patients under 18, the eligible visit note pool comprised 91,651 notes from 82,287 patients across 58 MGB psychiatry clinics (see Note Retrieval and Eligibility subsection of Supplement; Figure S1).

To support specificity to depression in the proposed analyses, our primary dataset (*MDD Cohort dataset*) included notes from patients with a lifetime (i.e. through Dec. 2021) diagnosis of major depressive disorder (MDD; ≥2 ICD codes), excluding patients with lifetime diagnoses of other major psychiatric conditions (obsessive-compulsive disorder [OCD], substance use disorder [SUD], bipolar disorder [BP], or schizophrenia [SCZ]; see Table S1 for diagnostic definitions). To test diagnostic specificity, a secondary dataset (*Diagnosis-Stratified Cohort dataset*) drew on these same diagnostic categories (and generalized anxiety disorder [GAD]) but required that each patient carry exactly one primary psychiatric diagnosis with no documented comorbidities

### Depression Severity Measures

Our analyses draw on four sources of depression severity information.

#### Patient-reported PHQ-9

PHQ-9 scores were retrieved from the RPDR for encounters during the 2015-2021 study period. Scores recorded within 7 days prior to an encounter were linked to that encounter; when multiple scores were available within this 7 day window, values were averaged.

#### Study-team clinician chart review

Two study-team clinicians (W.M., an attending psychiatrist, and Q.Y., a psychologist) independently assigned depression-specific CGI-S ratings to 125 notes from 30 patients sampled to span the full severity range (see Chart Review subsection of Supplement). Interrater reliability was assessed using quadratically weighted Cohen’s κ; because raters showed a systematic mean difference, we report both unadjusted and mean-adjusted κ ^23^.

#### Structured treating-clinician assessment

As an independent indicator of the treating clinician’s severity judgment, we identified the suicide risk assessment (SRA) — a structured rating adapted from the SAFE-T assessment ^21^ completed at every MGB outpatient psychiatric visit — as the most relevant routinely captured assessment (see Suicide Risk Assesment subsection of Supplement). Our analysis focused on a single item from this section: “What is the patient’s current, overall, acute risk of harm to self and/or others?” Responses are recorded on a four-point scale (1=minimal, 2=low, 3=moderate, 4=high). Prior work has demonstrated that this item shows good discrimination for suicide attempt within 90 days (outpatient AUC = 0.77) ^22^.

#### LLM-inferred scores

A large language model (OpenAI GPT-5.2) was provided with clinical notes via the HIPAA-compliant MGB Microsoft Azure infrastructure. Patient-reported outcome sections (including items and totals for the PHQ-9 and the correlated anxiety measure GAD-7) were redacted prior to inference to prevent information leakage (see Note Blinding subsection of Supplement). The model was prompted to infer three depression severity measures representing distinct assessment modalities: (1) PHQ-9 total score, a patient-reported, itemized measure ^24^; (2) Hamilton Depression Rating Scale (HAM-D) total score, a clinician-assessed, itemized measure ^25^; and (3) Clinical Global Impression-Severity (CGI-S), a clinician-assessed global severity rating ^26^. The CGI-S prompt was adapted from an anchored, transdiagnostic CGI-S framework ^27^ for use in the assessment of depressive symptoms (see LLM Prompting subsection of Supplement). Throughout, we refer to these as LLM PHQ-9, LLM HAM-D, and LLM CGI-S, respectively.

The model was prompted using the full-scale content of each measure. To assess whether results were sensitive to task specification, we tested multiple prompt formulations and model architectures (Table S2). Additional prompt formulations include (1) *Name-Only*, which included only the scale name, and (2) *Item-Level*, which required inference of each item response. For CGI-S, we also evaluated a shortened full-scale prompt (*Full-Scale Short*) to assess sensitivity to prompt detail and length (see LLM Prompting subsection of Supplement). The application of LLMs to clinical narrative notes was approved by the MGB Institutional Review Board (IRB#2018P002642 and IRB#2024P001444).

### Statistical Analysis

LLM-inferred depression severity scores were evaluated across four validity domains: convergent validity, predictive validity, diagnostic specificity, and consistency across clinics and demographic groups.

#### Convergent validity

We evaluated convergent validity by comparing LLM-inferred severity scores to three independent information sources: **(1) patient-reported depression severity** (PHQ-9), **(2) treating-clinician structured assessment** (SRA), and **(3) study-team clinician chart reviewer assessment** (depression-specific CGI-S). Agreement between LLM-inferred and patient-reported PHQ-9 scores (n=3,757 notes; 1,480 patients) and between LLM-inferred and study-team clinician CGI-S ratings (n=125 notes; 30 patients) was computed using quadratically weighted Cohen’s κ. Associations were further evaluated using Pearson correlations and repeated-measures correlations to account for within-patient dependence. For illustration, we compared LLM PHQ-9 with patient-reported PHQ-9 and LLM CGI-S with clinician-rated CGI-S using locally weighted scatterplot smoothing (LOWESS) in the Supplement. Lastly, AUCs were calculated for all severity scores in classifying two encounter-level clinical indicators: positive depression screen (patient-reported PHQ-9≥10, corresponding to moderate depression) and presence of suicide risk (clinician-assessed SRA≥2; i.e. greater-than-minimal suicide risk). The SRA analysis was restricted to notes with available SRA scores (n=2,985 notes; 653 patients). As a sensitivity analysis of the SRA AUC, the SRA portion of the note was redacted and AUC was recalculated (Note Blinding subsection of Supplement).

#### Predictive validity

We compared the ability of LLM-inferred scores and patient-reported PHQ-9 to predict two subsequent psychiatric events: **(1) antidepressant switching or augmentation**, defined as any medication change in which at least one new antidepressant was initiated relative to the prior regimen, including switches and additions (Table S3); and (**2) psychiatric emergency department (ED) visits**, defined as any ED encounter during which a psychiatry consult order was placed and a psychiatry progress note was signed. For all analyses, index time was defined as the date of the clinical visit at which either the patient-reported PHQ-9, clinician-assessed SRA, or LLM-inferred score was recorded. Patients were followed from index time until the outcome of interest and were censored at the end of follow-up (November 2021) or at their last recorded diagnosis. The number of eligible notes and events can be found in Table S4.

Kaplan–Meier curves compared time to anti-depressant switching among visits stratified by severity. For observed PHQ-9, we used standard cut points (<10 vs. ≥15). For each LLM-inferred score, visits were dichotomized at percentile cutoffs chosen to match the PHQ-9 group sizes (see Table S5). The same analytic approach was applied to observed SRA scores.

To account for multiple scores and events per patient, we used Andersen–Gill Cox proportional hazards models ^28^ implemented in R (R Foundation for Statistical Computation) using the “survival” package ^29^, allowing patients to contribute multiple risk intervals with variance clustered at the patient level. Scores were modeled continuously. To evaluate independent contributions, we fit joint models that included LLM-inferred scores together with either observed PHQ-9 or SRA scores. Discrimination was evaluated using Harrell’s concordance indices (C-index). Because the Andersen–Gill model assumes independence of recurrent events within individuals, we conducted a sensitivity analysis limited to the first event per patient to test robustness to this assumption.

#### Diagnostic specificity

Depression-threshold classification rates were compared across diagnostic groups in the *Diagnosis-Stratified Cohort Datase*t. For each diagnosis, analyses were restricted to notes explicitly documenting that diagnosis (maximum 500 notes per diagnosis). The proportion of encounters meeting moderate depression severity criteria (LLM PHQ-9≥10, LLM HAM-D≥14, LLM CGI-S≥4) was computed within each group. Higher classification rates in MDD relative to non-depression diagnoses indicate specificity. Confidence intervals were estimated using 1,000 bootstrap samples with replacement.

#### Consistency

To assess consistency across populations, we calculated the overall correlations between LLM-inferred and patient-reported PHQ-9 scores within demographic subgroups defined by age (18–64, 65+), gender (male, female), race (White, Black, Asian, Other), and ethnicity (Hispanic, non-Hispanic). Subgroup differences were evaluated relative to the largest category in each demographic category (e.g., 18-64 vs. 65+) using Fisher’s z-test.

Additionally, we tested whether the association between LLM-inferred and patient-reported PHQ-9 scores varied across clinics using a meta-analytic framework. Specifically, we conducted a heterogeneity analysis by allowing the slope (w) to vary by clinic:

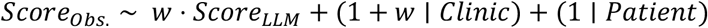

Between-clinic heterogeneity was quantified using the estimated between-clinic variance (τ^2^) and the proportion of total variance attributable to heterogeneity (I^2^). Analyses were conducted across 29 clinics for patient-reported PHQ-9 (n=3,757 notes; 1,480 patients) and 12 clinics for mean study clinician-rated CGI-S (n=125 notes; 30 patients) within the MGB system.

## Results

### Sample Characteristics and Score Distributions

The MDD analytic note dataset comprised 34,767 visit notes from 3,543 patients (Table 1; see Table S6 for demographic breakdown of *Diagnosis-Stratified Cohort Datase*t). Patient-reported PHQ-9 scores were available for 3,757 visit notes (10.8% of visits). Among notes with an observed PHQ-9, the median score was 9 (IQR: 4-14), corresponding to mild depression (Figure 1a). Median LLM-inferred scores were 7 for LLM PHQ-9, 10 for LLM HAM-D, 3 for LLM CGI-S, all corresponding to mild depression. Despite being prompted to infer scores for distinct measures, the three LLM-inferred scores were strongly intercorrelated (r>0.85; Figure 1b).

**Table 1:** Summary of demographic characteristics and psychometric data availability in primary MDD note dataset. Starting from all eligible notes (see Note Eligibility of Supplement; Figure S1), the cohort was restricted to individuals with lifetime diagnoses of major depressive disorder (MDD; ≥2 ICD at the end of the note draw) or generalized anxiety disorder (GAD), excluding those with a lifetime diagnoses of obsessive-compulsive disorder (OCD), substance use disorders (SUD), bipolar disorder (BP), or schizophrenia (SCZ). Patient-reported 9-item Patient Health Questionnaire (PHQ-9) scores were linked to a clinical note if completed within 7 days prior to the corresponding visit. Treating clinician–documented suicide risk assessments (SRA) were assigned if recorded on the same day as the visit. Values are presented as the number of notes and unique patients within each grouping. Percentages are shown relative to all eligible notes within the grouping and patients within each subgroup, except for SRA, where percentages reflect only the subset of notes from 2019 onward (SRA documentation was not widely implemented across Mass General Brigham until 2019). Cells containing fewer than 20 observations, along with the other subgroups in the same demographic category, were omitted to preserve patient privacy (denoted as ‘-’).

| Cat. | Group | All |  | w/ PHQ-9 |  | w/ SRA (since 2019) |  |
| --- | --- | --- | --- | --- | --- | --- | --- |
|  |  | Notes | Pat. | Notes | Patients | Notes | Patients |
| *All* |  | 34,767 | 3,543 | 3,757 (10.8%) | 1,480 (41.8%) | 2,985 (30.0%) | 653 (36.2%) |
| *Year* |  |  |  |  |  |  |  |
|  | 2015–2016 | 8,914 | 1,720 | 806 (9.0%) | 435 (25.3%) | NA | NA |
|  | 2017–2018 | 15,898 | 2,254 | 1,537 (9.7%) | 794 (35.2%) | NA | NA |
|  | 2019–2021 | 9,955 | 1,802 | 1,414 (14.2%) | 700 (38.8%) | 2,985 (30.0%) | 653 (36.2%) |
| *Age* |  |  |  |  |  |  |  |
|  | 18–64 | 26,583 | 2,709 | 3,111 (11.7%) | 1,194 (44.1%) | 2,242 (29.7%) | 462 (35.5%) |
|  | 65+ | 8,184 | 935 | 646 (7.9%) | 305 (32.6%) | 743 (30.9%) | 194 (37.8%) |
| *Gender* |  |  |  |  |  |  |  |
|  | Female | 23,471 | 2,516 | 2,708 (11.5%) | 1,081 (43.0%) | 1,838 (27.5%) | 438 (34.3%) |
|  | Male | 11,296 | 1,027 | 1,049 (9.3%) | 399 (38.9%) | 1,147 (35.1%) | 215 (40.9%) |
| *Race* |  |  |  |  |  |  |  |
|  | White | 28,940 | 2,867 | 3,073 (10.6%) | 1,205 (42.0%) | 2,493 (30.9%) | 560 (38.3%) |
|  | Black | 1,699 | 240 | 258 (15.2%) | 105 (43.8%) | 154 (25.5%) | 32 (26.4%) |
|  | Asian | 1,208 | 93 | 124 (10.3%) | 48 (51.6%) | 162 (37.2%) | 21 (42.9%) |
|  | Other | 2,161 | 221 | 196 (9.1%) | 85 (38.5%) | 143 (21.3%) | 26 (22.6%) |
| *Eth.* |  |  |  |  |  |  |  |
|  | Non-Hisp. | 33,581 | 3,364 | 3,635 (10.8%) | 1,434 (42.6%) | - | - |
|  | Hisp. | 1,186 | 179 | 122 (10.3%) | 46 (25.7%) | - | - |

**Figure 1:**
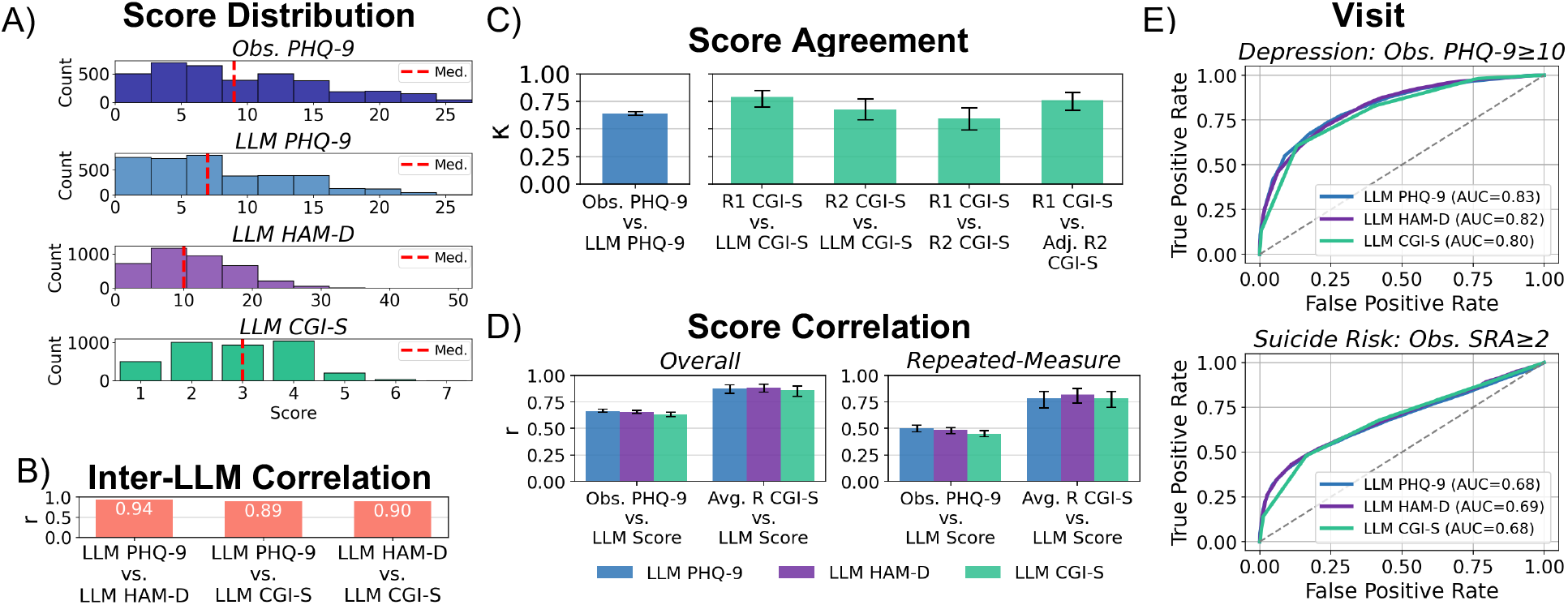
Distributions and convergent validity of LLM-inferred depression severity scores relative to established psychometrics. **A)** Distribution of depression severity scores among notes with an available patient-reported PHQ-9 in the MDD cohort dataset. Shown (from top to bottom): patient-reported PHQ-9, LLM-inferred PHQ-9, LLM-inferred HAM-D, and LLM-inferred CGI-S. Red dashed lines denote median scores. **B)** Pearson correlations (r) between LLM-inferred PHQ-9, HAM-D, and CGI-S scores. **C)** Score agreement between LLM-inferred and patient-reported PHQ-9 scores, and between two study-staff clinician CGI-S ratings. Agreement was assessed using Cohen’s kappa (k) with quadratic weighting. Rater 2 scores were additionally mean adjusted to match rater 1 scores (see Chart Review subsection in Supplement) **D)** The overall and repeated measure correlations between LLM-inferred PHQ-9, HAM-D, and CGI-S scores and both patient-reported PHQ-9 and the average study clinician-rated CGI-S (Avg. R CGI-S). **C)** Receiver operating characteristic (ROC) curves and areas under the curve (AUCs) for classifying moderate-to-severe depression (Obs. PHQ-9≥10) and clinician-assessed greater-than-minimal suicide risk (SRA≥2) at each visit based on LLM-inferred depression severity scores. Supplemental Figure S3 presents a sensitivity analysis for the SRA≥2 outcome, showing the AUC after redacting the SRA section from the notes to evaluate potential information leakage. All analyses were conducted within the MDD cohort, and 95% confidence intervals are shown where available (except for panel B as confidence intervals were negligible).

### Convergent Validity

#### Convergence with patient-reported PHQ-9

Agreement between the LLM PHQ-9 with patient-reported PHQ-9 was moderate (κ=0.64, 95%CI: 0.62–0.66; r=0.67, 95%CI: 0.65–0.68), and attenuated after accounting for within-patient dependence (repeated-measures r=0.50; 95%CI: 0.47–0.53). LLM PHQ-9 achieved an AUC of 0.83 (95%CI: 0.81–0.84) for classifying a positive depression screen (Obs. PHQ-9≥10). A LOWESS curve indicated an approximately linear relationship between estimated and patient-reported PHQ-9s, although the association deviated from a one-to-one correspondence (Figure S2).

#### Convergence with treating-clinician assessment

LLM CGI-S also showed moderate convergence with the treating-clinician’s structured assessment, achieving an AUC of 0.69 (95%CI: 0.67–0.71) for classifying presence of suicide risk (SRA≥2; greater-than-minima” suicide risk). AUCs were unchanged when performing the same analysis with the SRA portion of the notes redacted (see Figure S3).

#### Convergence with chart review

LLM-inferred scores correlated strongly with study-team clinician chart review (LLM CGI-S vs. mean chart-review CGI-S: r=0.86, 95%CI: 0.80–0.90), and this association remained strong after accounting for within-patient dependence (r=0.79, 95%CI: 0.70–0.85). LLM CGI-S showed higher interrater agreement with each clinician rater (κ=0.79 with Rater 1, 95%CI: 0.70–0.85; κ=0.67 with Rater 2, 95%CI: 0.58–0.77) than the raters showed with each other (κ=0.59, 95%CI: 0.49–0.69; adjusted κ=0.76 after correcting for a systematic mean difference, 95%CI: 0.67–0.8; see Chart Review subsection in Supplement).

### Predictive Validity

We compared the ability of LLM-inferred scores, patient-reported PHQ-9, and treating-clinician SRA to predict subsequent antidepressant switching and psychiatric ED visits.

For antidepressant switching, Kaplan–Meier survival curves stratified by LLM-inferred severity closely paralleled those derived from observed PHQ-9 and SRA (Figure 3a; Figure S6). Cox models showed modest discrimination across all measures (C-index≈0.60; Figure 3b; Tables S7–S9). Moderate performance may reflect switching driven by factors unrelated to depression severity, such as treatment side effects.

For future psychiatric ED visits, LLM-inferred scores showed slightly stronger predictive discrimination (C-index≈0.65). Among visit notes with an available patient-reported PHQ-9, each one-point increase in LLM CGI-S was associated with a 45% higher hazard of psychiatric ED visit (95%CI: 21%-74%), with discrimination (C-Index) of 0.63 (95%CI: 0.57–0.68). In the same subset of notes, patient-reported PHQ-9 showed minimal improvement over LLM CGI-S in discrimination (C-index=0.65; 95%CI: 0.60–0.71) with the same set of notes. Jointly modeling LLM CGI-S and PHQ-9 did not meaningfully improve discrimination (C-index = 0.66; 95%CI: 0.60–0.71; Table S9). Among notes with clinician-assessed SRA, predictive performance for psychiatric ED visits was similar (C-index=0.61; 95%CI: 0.51–0.71). Results were largely consistent when analyses were limited to first events only (Table S7-S9).

### Diagnostic Specificity

In the *Diagnosis-Stratified Cohort Dataset*, LLM-inferred scores showed clear specificity to depression. Among notes from patients with MDD as their sole psychiatric diagnosis, 40.0% (95%CI: 38.1-42.0) were classified as at least moderately depressed (LLM CGI-S ≥4), compared with fewer than 10% of notes for patients with only GAD, OCD, SUD, or schizophrenia (Figure 2). Patients with bipolar disorder had 17.0% (95%CI: 13.5-21.1) of notes classified as at least moderately depressed, and the fact that bipolar disorder had a higher LLM classification of moderate depression compared to the other comparator groups is consistent with the high prevalence of depressive symptoms in bipolar disorder ^30^.

**Figure 2.**
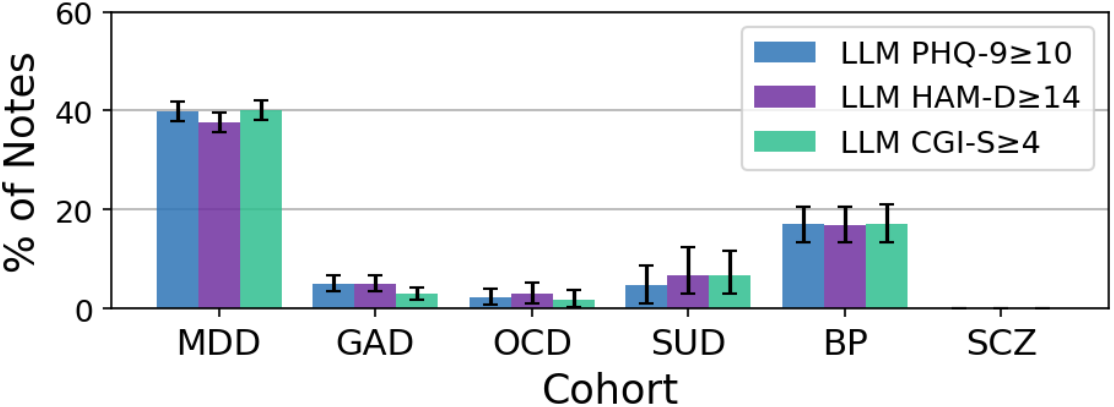
Discriminant validity of LLM-inferred depression severity across psychiatric diagnoses. Percentage of clinical notes classified as at least moderately depressed (LLM PHQ-9 ≥10, LLM HAM-D ≥14, or LLM CGI-S ≥4) within a diagnosis-stratified cohort (MDD, GAD, OCD, SUD, BP, SCZ). Error bars indicate 95% confidence intervals derived from 1,000 bootstrap resamples.

**Figure 3:**
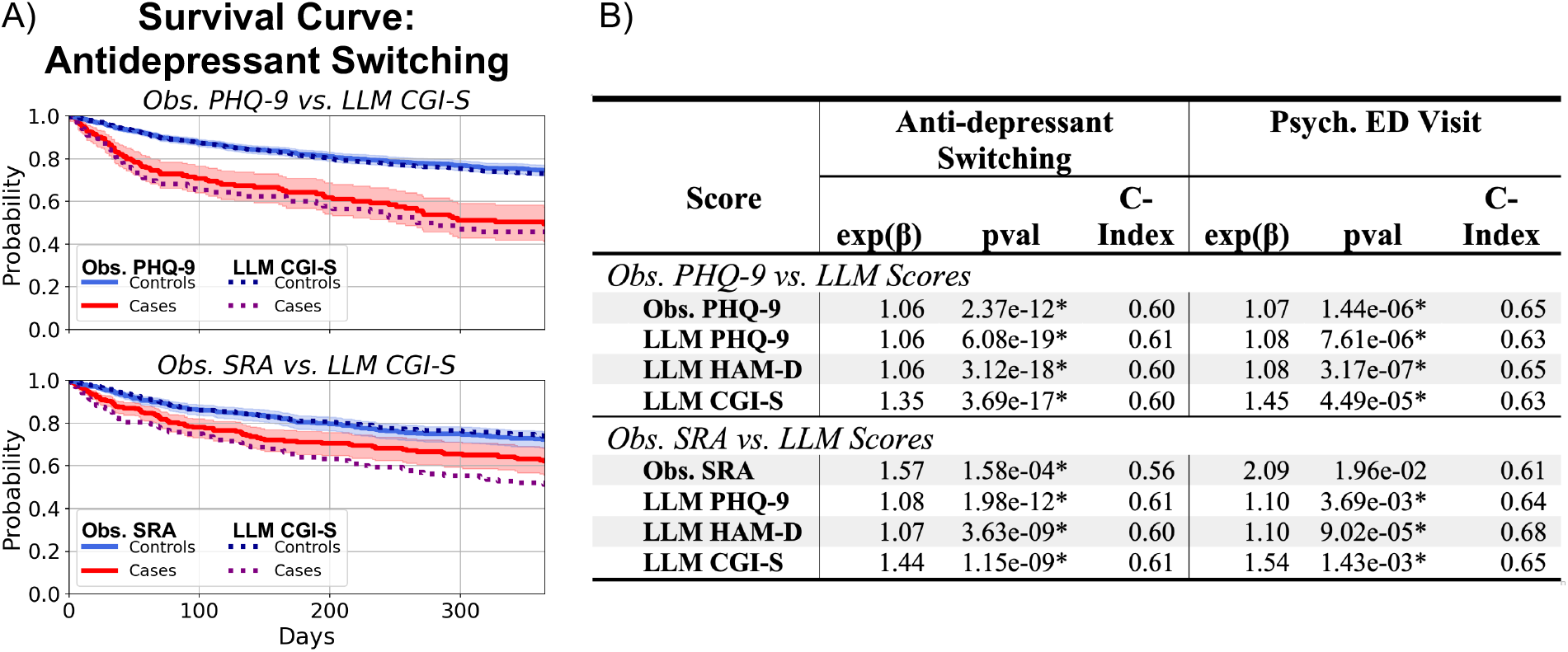
Predictive validity of LLM-inferred depression severity for antidepressant switching and psychiatric emergency visits. **A)** Case-control Kaplan-Meier survival curves for time to antidepressant switching or augmentation were generated to compare risk stratification using: (1) patient-reported PHQ-9 scores (cases ≥20; controls <10) versus LLM-inferred GCI-S scores (cases ≥5; controls <4), and (2) clinician-rated suicide risk assessment (cases ≥2; controls <2) versus LLM-inferred CGI-S scores (cases ≥4 controls <4). Shaded regions denote 95% confidence intervals for observed PHQ-9 and suicide risk assessment case-control curves. Corresponding percentage cutoffs for each definition are reported in Supplemental Table S5. **B)** Results from Andersen-Gill Cox proportional hazards models estimating the risk of antidepressant switching and future psychiatric emergency department (i.e. psych ED) visits based on patient-reported PHQ-9, clinician-assessed SRA, and LLM CGI-S. Models were conducted separately in two datasets: notes containing a patient-reported PHQ-9 and notes containing a clinician-assessed SRA. All models were clustered by patient to account for repeated measures and outcomes (pval<.05/8 *). Model discrimination for both outcomes was assessed using the concordance index (C-index). 95% confidence intervals and sensitivity analyses for all three LLM-inferred scores can be found in Table S7-S9.

### Consistency

Across demographic strata (Table 2), the correlation between LLM CGI-S and patient-reported PHQ-9 scores was generally moderate (e.g., r=0.62 for females and r=0.65 for males). Correlations were lower among Black patients than White patients (r=0.48 vs. 0.64; p<0.001*) and lower among Hispanic patients than among non-Hispanic patients (r=0.43 vs. 0.63; p=0.002*). Correlations were also slightly lower among patients aged ≥65 years compared with those aged 18–64 (r=0.58 vs. 0.64; p=0.028), and among patients in the “Other” race category compared with White patients (r=0.54 vs. 0.64; p=0.037); however, these differences were not statistically significant after correction for multiple testing.

**Table 2.** Consistency of agreement across demographic groups. For subgroups defined by age, gender, race, and ethnicity, the table reports the correlation between LLM-inferred CGI-S scores and patient-reported PHQ-9 scores, along with 95% confidence intervals. P-values are calculated using Fisher’s z transform to compare each subgroup’s correlation with that of the reference group within the same demographic category (pval<.05/6 *).

| *Obs. PHQ-9 vs. LLM CGI-S* |  |  |  |  |
| --- | --- | --- | --- | --- |
| Cat. | Group | r | ref. Group | pval |
| *Age* |  |  |  |  |
|  | 18-64 | 0.64 (0.61-0.66) |  |  |
|  | 65+ | 0.58 (0.52-0.63) | 18-64 | 0.028 |
| *Gender* |  |  |  |  |
|  | Female | 0.62 (0.59-0.64) |  |  |
|  | Male | 0.65 (0.62-0.69) | Female | 0.103 |
| *Race* |  |  |  |  |
|  | White | 0.64 (0.62-0.66) |  |  |
|  | Black | 0.48 (0.38-0.57) | White | <0.001* |
|  | Asian | 0.70 (0.59-0.78) | White | 0.297 |
|  | Other | 0.54 (0.43-0.63) | White | 0.037 |
| *Ethn.* |  |  |  |  |
|  | Non-Hisp. | 0.63 (0.61-0.65) |  |  |
|  | Hisp. | 0.43 (0.27-0.56) | Non-Hisp. | 0.002* |

Associations between LLM-inferred and patient-reported PHQ-9 and the mean study clinician-rated CGI-S were consistent across MGB clinics, with low between-clinic heterogeneity (I^2^<0.1; Table S10).

### Sensitivity Analyses

Findings were qualitatively unchanged across other recent OpenAI GPT models (i.e., GPT-4o-mini, GPT-4o, GPT-5-mini, GPT-5, GPT-5.2; Supplemental Table S11; Figure S5a). Results were also robust to the prompting target (PHQ-9, HAM-D, and depression-specific CGI-S) and prompting format (scale name only vs full instrument text; item-level inference with post hoc summation where applicable) (see Figure S5b and S6).

## Discussion

There is increasing interest in leveraging real-world data from EHRs for psychiatric research, including pharmacoepidemiology, risk stratification, and treatment effectiveness studies ^4–6^. Many of these applications are limited, however, by the fact that symptom severity and treatment response are poorly captured in structured EHR fields. In this study, we subjected LLM-inferred depression severity scores from psychiatric outpatient notes to a comprehensive validation battery. LLM-inferred scores showed strong convergence with clinician chart review (r=0.86), moderate agreement with patient-reported PHQ-9 (r=0.63), good classification accuracy for positive depression screening (AUC=0.80 for PHQ-9≥10), specificity to depression over other psychiatric conditions, and predictive validity for subsequent psychiatric events. For predicting psychiatric ED visits, LLM-inferred scores performed similarly to both patient-reported PHQ-9 scores and treating-clinician overall (single-item) suicide risk assessments. Together, these results support LLM-inferred note-based severity as a valid measure.

The pattern of stronger convergence with clinician judgment than with patient-reported PHQ-9 is consistent with prior work reporting high LLM–chart-reviewer agreement at another institution ^19^ and more modest LLM–PHQ-9 concordance ^18^. Our results help clarify this discrepancy. The predictive validity findings argue against a purely artifactual explanation: if LLM scores and chart reviewers were simply converging on documentation habits rather than clinical severity, LLM-inferred scores would not perform similarly as PHQ-9 in predicting independent downstream events. The fact that LLM scores predict future events as well as PHQ-9 scores despite being only moderately correlated with PHQ-9 scores could reflect the possibility that clinical notes may encode dimensions of severity that individual measures do not fully capture, including functional impairment, mental status observations, safety concerns, and the treating clinician’s integrated judgment. Sensitivity analyses across prompt formulations and model architectures further indicate that the discrepancy is not an artifact of task specification. More broadly, LLM-inferred PHQ-9, HAM-D, and CGI-S scores were strongly intercorrelated despite representing distinct assessment modalities, and results were largely stable across prompting strategies — suggesting that the LLM is extracting a common severity signal from the notes rather than performing differently depending on the specific measure it is asked to infer.

Convergent validity also varied modestly by race and ethnicity, with lower PHQ-9–LLM CGI-S correlations among Black vs White and Hispanic vs non-Hispanic patients. Prior work suggests the PHQ-9 does not exhibit differential item functioning across racial and ethnic groups ^31^, pointing instead to differences in what is elicited and documented in clinical notes ^32^ and variation in PHQ-9 administration and availability as more likely sources of measurement heterogeneity. True differences in symptom presentation or in the relationship between patient-reported and clinician-rated severity across groups may also play a role. Additionally, demographic cues in clinical documentation could influence LLM-inferred severity ^33^. Disentangling these mechanisms and ensuring equitable model performance across populations is a critical priority for future multi-site validation studies.

These findings have different implications depending on PHQ-9 availability. For the majority ^7^ of psychiatric visits without standardized symptom measures, LLM-inferred severity scores may help recover depression severity information from routine clinical documentation. In visits where PHQ-9 scores were already available, however, we did not observe clear evidence of added value in this analysis, though this may reflect limited power or the specific parameterization used. If externally validated across settings and demographic groups, these scores could support more complete longitudinal severity tracking for retrospective research, quality improvement, and measurement-based care.

Several limitations should be noted. This study was conducted within a single healthcare system (MGB), though the consistency of LLM–chart-review agreement across our clinics (I^2^<0.10) and similar findings at Johns Hopkins ^19^ provide preliminary cross-site evidence; formal multi-site validation using a shared protocol remains needed. The chart-review sample was modest (125 notes from 30 patients), and raters did not achieve full calibration, necessitating adjustment for a systematic mean difference. The SRA, while the most relevant structured assessment available, captures suicide risk rather than depression severity per se and is an imperfect proxy for the treating clinician’s overall severity judgment. Patient-reported PHQ-9 scores were missing non-randomly, likely reflecting clinical workflow as much as patient state, which may bias the convergent validity comparison. Our cohort was restricted to psychiatric outpatient settings; generalizability to primary care, where the majority of depression is treated ^34^ and documentation practices differ substantially, has not been established. The diagnostic specificity analysis used patients with a single psychiatric diagnosis, who may not represent the comorbid presentations typical of routine practice. Finally, we did not evaluate performance across clinician credentials — the last of which may be particularly important given that documentation practices and clinical assessments likely vary across provider types ^22^ .

The present findings provide initial validity evidence for LLM-inferred depression severity scores from routine EHR documentation, especially for extending severity measurement to visits where standardized measures were not collected. Broader use will require multi-site replication, external validation across settings and demographic groups, and careful attention to score presentation and guardrails for appropriate use. If these steps are successful, LLM-informed severity tracking could improve EHR-based research and, ultimately, clinical care.

## Supporting information

LLM Prompting

Scale Text

Chart Review Instructions

## Data Availability

Due to the presence of Protected Health Information (PHI), access to the clinical data used in this study is restricted. The data were obtained under Massachusetts General Brigham Institutional Review Board approval for use only in the current study. As a result, the dataset is not publicly available.

## Acknowledgements

We thank Dr. Kate H. Bentley for the facilitation of the collection of the suicide risk assessment data within Mass General Brigham. MC is supported by the MGB Training Program in Precision and Genomic Medicine (T32HG010464). WM is supported by the National Library of Medicine/NIH grant T15LM007092. BW is supported in part by funding from the Brain and Behavior Research Foundation Young Investigator Award and NIMH grant P50MH129699. CK was supported in part by NIMH K01 MH135131. JWS is supported in part by R01 MH137218.

