## Supplemental Material for "Multi-Criteria Validation of LLM-Inferred Depression Severity from Outpatient Psychiatry Notes"

### Supplemental Methods

#### Note Eligibility

Using the MGB Research Patient Data Registry (RPDR), we acquired all narrative clinical notes for all 95,157 patients enrolled in the Massachusetts General Brigham (MGB) Biobank as of November 2021, as well as for 100,000 randomly sampled patients not enrolled in the Biobank [20]. The notes spanned from March 1976 (earliest available) to November 2021 (date of acquisition) and comprised 11 note types: cardiology reports, discharge summaries, endoscopy reports, operative notes, pathology reports, pulmonary notes, radiology reports, Epic history and physical notes, progress notes, ambulatory visit notes, and Longitudinal Medical Records (LMR) visit notes.

To ensure consistency following MGB's transition to Epic, we restricted the dataset to notes dated April 2015 or later. During preprocessing, notes written by the same provider for the same patient on the same day were concatenated.

For this study, we focused specifically on outpatient psychiatric visit notes. Psychiatric departments were identified using structured clinic name data available in RPDR or by detecting the term "Psych" in department names within the clinical notes. After applying these criteria, the final outpatient psychiatric clinical note dataset included 118,711 visit notes from 9,066 patients across 71 MGB psychiatric clinics.

We then applied additional exclusion criteria. We removed group therapy notes, MGB-censored sensitive notes (i.e., notes for patients with protected status), notes authored by non-clinician providers (those without MD, DO, PhD, PsyD, PA, NP, or CNP credentials), and notes from visits involving patients younger than 18 years of age. The resulting note pool was subsequently sampled to construct the *MDD Cohort dataset* and the *Diagnosis-Stratified Cohort dataset (*See Figure S1a).

#### Suicide Risk Assessment

The MGB suicide risk assessment (SRA) is a structured electronic health record (EHR) form (flowsheet) based on the Substance Abuse and Mental Health Services Administration's validated Suicide Assessment Five-Step Evaluation and Triage (SAFE-T) resource. Per Joint Commission requirements, treating clinicians complete the SRA at every psychiatric visit within MGB, with widespread adoption beginning after 2019. The assessment evaluates recent suicidal thoughts and behaviors, nonsuicidal self-injury, and violent or destructive thoughts or actions, and includes items assessing suicide risk factors (e.g., depressed mood, recent loss, access to firearms) and protective factors (e.g., social support). The assessment concludes with the question, "What is the patient's current, overall, acute risk of harm to self and/or others?" with response options of 1=minimal, 2=low, 3=moderate, or 4=high. The full item-level SRA is provided in Supplement Table 1 of Bentley et al [22].

SRAs were obtained from MGB's Enterprise Data Warehouse (Snowflake) and matched to corresponding clinical notes using patient identification numbers and note dates.

### Note Blinding

#### *Note preprocessing*

Because notes were transferred into text files, minor artifacts were introduced. Prior to blinding, notes were minimally cleaned by removing the first two header lines, stripping page-break artifacts, and deleting residual character artifacts.

#### *PHQ-9 and GAD-7 Blinding*

Clinical notes contain free-text clinical narratives as well as structured tabular data (e.g., immunizations, allergies, medications). They also can include PROMs data for the visit, including PHQ-9 and GAD-7 scores from the current and prior encounters. Because the goal was to evaluate the LLM's ability to *infer,* not *extract,* depression severity from clinical narrative, all PHQ-9 and GAD-7 item-level responses, summary scores, and severity descriptors were removed. Blinding was performed using a deterministic, rule-based text redaction pipeline:

1. **PROMs section excision:** Text was first segmented into lines and scanned for known PROMs headers. When a PROMs header was detected, the entire PROMs block was removed and replaced with a fixed placeholder (`[REDACTED PROMS SECTION]`). The redaction continued until a predefined non-PROMs clinical section header was encountered, ensuring complete removal of questionnaire content without affecting later sections of the note. This step was applied recursively to ensure complete removal of all PROMs sections.
2. **Question-level and score redaction:** The remaining text was searched for PHQ-9 and GAD-7 question content using case-insensitive and punctuation-agnostic matching. When a question was identified, the full contiguous block containing the question and its associated response(s) was replaced with `[REDACTED]`.
3. **Score and severity masking:** Any sentence containing PROMs-related terms (e.g., "PHQ", "GAD", "total", "PROMs") was masked by replacing all numeric values and standardized severity descriptors (e.g., "none", "minimal", "mild", "moderate", "severe", "depression", "anxiety") with a non-informative placeholder (`?`).

Staff scientists (MC) randomly reviewed 100 notes to verify that PROMs data had been adequately redacted. Additionally, during rating, expert clinicians also flagged any residual information leakage. Among the 125 notes that were rated, no major leaks (i.e., sufficient information to reconstruct the total PHQ-9 or GAD-7 scores) were observed. However, partial leaks, consisting of minor isolated score components, were identified in 4% of rated notes according to the study psychiatrist and 8.8% of rated notes according to the study psychologist. The difference in reported rates of partial leakage likely reflects the difficulty of detecting isolated score components within large volumes of unstructured clinical text.

#### *SRA Blinding*

As a sensitivity analysis, suicide risk assessment (SRA) content was fully blinded to prevent direct disclosure of suicide risk evaluations (this blinding was not applied in the primary analyses).

- **Identification:** Notes were segmented into lines and scanned for the SRA section header ("Suicide Risk Assessment") using case-insensitive matching. After a header was detected, subsequent lines were examined for item-level SRA question text using keyword-based matching (e.g., references to suicidal or homicidal thoughts, self-harm, violence, risk factors, or safety planning). The SRA section was defined to span from the header through the final line containing such question text. To reduce false positives, candidate SRA sections longer than 150 lines were excluded.
- **Redaction:** Identified SRA sections were fully removed and replaced with a fixed placeholder [REDACTED SRA SECTION]. Notes without a detected SRA section were left unchanged.

Staff scientist (MC) randomly reviewed 100 notes containing SRA content to confirm that SRA sections were correctly and completely blinded.

#### LLM Prompting

LLMs were prompted using a system prompt, a user prompt, and a JSON schema to constrain model outputs to integers within a provided range. In addition to the primary full-scale (*Full-Scale*) prompting approach, we evaluated two alternative prompting strategies: 1) *Name-Only*, which prompted the LLM to infer scores using only the scale name and 2) *Item-Level*, which prompted the LLM to infer each questionnaire item using its full text. For LLM-based CGI-S scoring, we also evaluated a shortened version of the full-scale prompt (*Full-Scale Short*) to assess sensitivity to prompt length. Due to prompt length (including JSON schema) and structural complexity, full prompt specifications and scale texts are provided in *Supp_LLM_Prompting.pdf* and *Supp_Scale_Text.pdf*.

All analyses were restricted to OpenAI models due to existing agreements enabling HIPAA-compliant deployment within the MGB Microsoft Azure infrastructure. Therefore, the models tested were gpt-4o-mini, gpt-4o, gpt-5-mini, gpt-5, and gpt-5.2. Temperature was set to 0 for all models to ensure deterministic outputs where possible (i.e. gpt-4o, gpt-4o-mini). See Supplemental Table S2 for more details on GPT parameters used.

#### Chart Review

An attending psychiatrist (WM) and a psychologist (QY) independently rated depression severity for 125 outpatient psychiatric visit notes. Ratings were assigned using a depression-specific Clinical Global Impression-Severity (CGI-S) scale, based on clinical judgment and the study's predefined scoring rules. The depression-specific CGI-S was adapted from Dunlop (2017) by modifying the anchor descriptions to refer specifically to "depressive symptoms" rather than general "symptoms."

Raters followed standardized written instructions (see *Supp_Chart_Review_Instructions.pdf* for the full protocol). Two calibration rounds were conducted before the final chart review (Round 1: 12 notes from 3 patients; Round 2: 22 notes from 5 patients). Discrepancies were discussed after each round to clarify and refine the rating guidelines. Following calibration, the raters reached consensus with each other and the lead author (MC) on the final scoring approach.

The final sample included 125 notes from 30 patients (intake plus 3–5 follow-up visits per patient). To ensure representation across the full range of depression severity, 20 patients were randomly selected based on their initial patient-reported PHQ-9 scores, with four patients drawn from each severity category (None–Minimal, Mild, Moderate, Moderately Severe, Severe). An additional 10 patients without documented patient-reported PHQ-9 scores were randomly selected to reduce potential bias related to score availability in the clinical notes.

Inter-rater reliability (IRR) was assessed using quadratically weighted Cohen's κ. Because full calibration (IRR>0.8) was not achieved after calibration rounds, we report both unadjusted and adjusted IRR. Adjustment accounted for mean rater differences by subtracting the mean score difference (1.05 rounded to 1; see *Results*) from the higher-scoring rater and clipping scores to the 1–7 range (i.e., $Adj.CGIS_{R2} = CGIS_{R2} - 1, bounded\ between\ 1\ to\ 7$).

### Supplemental Tables:

**Supplemental Table S1. Psychiatric diagnosis codes.** Lists of ICD-9 and ICD-10 diagnostic codes used to identify psychiatric conditions examined in this study, including major depressive disorder (MDD), generalized anxiety disorder (GAD), obsessive-compulsive disorder (OCD), substance use disorder (SUD), bipolar disorder (BP), and schizophrenia (SCZ). Lifetime diagnosis was assigned if a patient ≥2 ICD codes corresponding to a diagnosis.

| Dx | ICD 9 Codes | ICD 10 Codes |
|---|---|---|
| **MDD** | 296.2x; 296.3x | F32.x-33.x |
| **GAD** | 300.02 | F41.1 |
| **OCD** | 300.3 | F42.x |
| **SUD** | 303.x; 304.x; 305.x (*excluding 305.1*) | F10-F16; F18-F19 (*excluding F1x.90*) |
| **BP** | 296.0x; 296.1x; 296.4x–296.8x (*excluding 296.82*) | F30.x; F31.x |
| **SCZ** | 295.x | F20.x |

**Supplemental Table S2. OpenAI GPT model details.** A summary of the OpenAI GPT models evaluated in this study, including model names, version release dates, temperature configurations, content filter settings, and input/output pricing per 1 million tokens. Temperature parameters were not applicable for GPT-5 and later models because these models use internal reasoning mechanisms that do not expose user-adjustable sampling controls. Only OpenAI models were evaluated, consistent with existing agreements that enable HIPAA-compliant deployment within the MGB Microsoft Azure infrastructure. Pricing information was obtained from the Azure OpenAI Service Website as of February 19, 2026.

| OpenAI Model | Misc. | | Filter Settings | | | | Cost | |
|---|---|---|---|---|---|---|---|---|
| | Version Date | Temp. | Violence | Hate | Sexual | Self-Harm | Input Pricing (1M Tokens) | Output Pricing (1M Tokens) |
| **GPT-4o-mini** | 2024-07-18 | 0 | Lowest | Lowest | Lowest | Lowest | $0.15 | $0.60 |
| **GPT-4o** | 2024-11-20 | 0 | Lowest | Lowest | Lowest | Lowest | $2.50 | $10 |
| **GPT-5-mini** | 2025-08-07 | NA | Lowest | Lowest | Lowest | Lowest | $0.25 | $2 |
| **GPT-5** | 2025-08-07 | NA | Lowest | Lowest | Lowest | Lowest | $1.25 | $10 |
| **GPT-5.2** | 2025-12-11 | NA | Lowest | Lowest | Lowest | Lowest | $1.75 | $14 |

**Supplemental Table S3. List of antidepressant medication names**. All medications listed below were used to identify antidepressant switching or augmentation events. These events were defined as any medication change in which at least one new antidepressant was initiated relative to the prior regimen, including both switches and additions.

| Name | Class |
|---|---|
| **Citalopram** | SSRI (Selective Serotonin Reuptake Inhibitor) |
| **Escitalopram** | SSRI (Selective Serotonin Reuptake Inhibitor) |
| **Fluoxetine** | SSRI (Selective Serotonin Reuptake Inhibitor) |
| **Fluvoxamine** | SSRI (Selective Serotonin Reuptake Inhibitor) |
| **Paroxetine** | SSRI (Selective Serotonin Reuptake Inhibitor) |
| **Sertraline** | SSRI (Selective Serotonin Reuptake Inhibitor) |
| **Vortioxetine** | SMS (Serotonin Modulator and Stimulator; multimodal antidepressant) |
| **Vilazodone** | SPARI (Serotonin Partial Agonist-Reuptake Inhibitor) |
| **Duloxetine** | SNRI (Serotonin-Norepinephrine Reuptake Inhibitor) |
| **Venlafaxine** | SNRI (Serotonin-Norepinephrine Reuptake Inhibitor) |
| **Desvenlafaxine** | SNRI (Serotonin-Norepinephrine Reuptake Inhibitor) |
| **Levomilnacipran** | SNRI (Serotonin-Norepinephrine Reuptake Inhibitor) |
| **Mirtazapine** | NaSSA (Noradrenergic and Specific Serotonergic Antidepressant) |
| **Bupropion** | NDRI (Norepinephrine-Dopamine Reuptake Inhibitor) |
| **Amitriptyline** | TCA (Tricyclic Antidepressant) |
| **Imipramine** | TCA (Tricyclic Antidepressant) |
| **Desipramine** | TCA (Tricyclic Antidepressant) |
| **Trimipramine** | TCA (Tricyclic Antidepressant) |
| **Clomipramine** | TCA (Tricyclic Antidepressant) |
| **Maprotiline** | TeCA (Tetracyclic Antidepressant) |
| **Doxepin** | TCA (Tricyclic Antidepressant) |
| **Nomifensine** | NDRI (Norepinephrine-Dopamine Reuptake Inhibitor) |
| **Phenelzine** | MAOI (Monoamine Oxidase Inhibitor) |
| **Moclobemide** | RIMA (Reversible MAO-A Inhibitor) |
| **Selegiline** | MAOI (MAO-B Inhibitor) |
| **Tranylcypromine** | MAOI (Monoamine Oxidase Inhibitor) |
| **Isocarboxazid** | MAOI (Monoamine Oxidase Inhibitor) |

**Supplemental Table S4. Eligible events and patients with antidepressant switching or psychiatric ED visits.** The number of events and unique patients identified between 2015 and 2021 with either (1) *antidepressant switching or augmentation* or (2) *psychiatry emergency department (ED) visits.* Events were considered eligible if they occurred after the first available clinical note and before censoring. Counts are shown for notes with an available patient-reported PHQ-9 and notes with an available clinician-assessed SRA after applying censoring. Patients were censored at the end of follow-up (November 2021) or at their last recorded diagnosis.

| | Anti-depressant Switching | | | | Psych. ED Visit | | | |
|---|---|---|---|---|---|---|---|---|
| | # of Events | # of Patients w/ Event | # of Notes | # of Patients | # of Events | # of Patients w/ Event | # of Notes | # of Patients |
| ***Obs. PHQ-9*** | 953 | 686 | 3,279 | 1,367 | 107 | 97 | 3,707 | 1,479 |
| ***Obs. SRA*** | 370 | 260 | 2,795 | 638 | 33 | 29 | 2,939 | 653 |

**Supplemental Table S5. Case-control thresholds and corresponding percentages used for survival curve analyses.** Thresholds were defined using established clinical cutoffs. For patient-reported PHQ-9 scores, controls were defined as <10 (no to minimal depression) and cases as ≥20 (severe depression). For clinician-assessed SRA, controls were defined as a score of 1 (no risk) and cases as scores ≥2 (greater-than-minimal risk). Percentiles were then derived from these cutoffs, and LLM-inferred depression score thresholds were selected to match as closely as possible the corresponding percentiles observed for PHQ-9 and SRA.

| | Control | | Case | |
|---|---|---|---|---|
| **Score** | **Thres.** | **%** | **Thres.** | **%** |
| *Obs. PHQ-9 vs. LLM Scores* | | | | |
| **Obs. PHQ-9** | <10 | 56.66 | ≥20 | 8.08 |
| **LLM PHQ-9** | <8 | 56.36 | ≥17 | 7.72 |
| **LLM HAM-D** | <12 | 57.49 | ≥20 | 8.45 |
| **LLM CGI-S** | <4 | 68.22 | ≥5 | 5.92 |
| *Obs. SRA vs. LLM Scores* | | | | |
| **Obs. SRA** | <2 | 68.37 | ≥2 | 31.63 |
| **LLM PHQ-9** | <8 | 65.44 | ≥8 | 34.56 |
| **LLM HAM-D** | <13 | 70.91 | ≥13 | 29.09 |
| **LLM CGI-S** | <4 | 74.49 | ≥4 | 25.51 |

**Supplemental Table S6: Demographic characteristics of the clinical notes and patients in the *Diagnosis-Stratified Cohort Dataset***. Demographic categories include age, gender, race, and ethnicity. The table also shows demographic distributions for notes with any diagnosis and for notes restricted to specific diagnoses: major depressive disorder (MDD), generalized anxiety disorder (GAD), obsessive compulsive disorder (OCD), substance use disorder (SUD), bipolar disorder (BP), or schizophrenia (SCZ). For each note diagnosis category, the table reports the number of clinics represented by all notes and patients with the available scores. To protect patient privacy, if any group within a demographic category had fewer than 20 patients for a given diagnosis, the entire category was omitted for that diagnosis to prevent counts from being inferred from the remaining totals. For OCD, SUD, and SCZ cohorts, the total patient counts were additionally reported as <50 to protect patient privacy.

| DX | # of | All | Age | | Gender | | Race | | | | Ethnicity | |
|---|---|---|---|---|---|---|---|---|---|---|---|---|
| | | | 40–64 | 65+ | Fem. | Male | White | Black | Asian | Other | Non-Hisp. | Hisp. |
| ***Any*** | *Notes* | 13,805 | 10,517 | 3,084 | 9,199 | 4,606 | 11,230 | 829 | 511 | 872 | 13,204 | 601 |
| | *Patients* | 1,767 | 1,324 | 450 | 1,191 | 576 | 1,419 | 132 | 49 | 108 | 1,676 | 91 |
| ***MDD*** | *Notes* | 9,988 | 7,362 | 2,477 | 6,735 | 3,253 | 7,741 | 713 | 408 | 796 | 9,420 | 568 |
| | *Patients* | 1,292 | 942 | 355 | 894 | 398 | 1,002 | 113 | 31 | 94 | 1,208 | 84 |
| ***GAD*** | *Notes* | 1,714 | 1,450 | 262 | 1,195 | 519 | | | | | | |
| | *Patients* | 242 | 199 | 43 | 171 | 71 | | | | | | |
| ***OCD*** | *Notes* | 277 | | | | | | | | | | |
| | *Patients* | <50 | | | | | | | | | | |
| ***SUD*** | *Notes* | 104 | | | | | | | | | | |
| | *Patients* | <50 | | | | | | | | | | |
| ***BP*** | *Notes* | 1,668 | 1,362 | 265 | 1,048 | 620 | | | | | | |
| | *Patients* | 145 | 112 | 35 | 86 | 59 | | | | | | |
| ***SCZ*** | *Notes* | 54 | | | | | | | | | | |
| | *Patients* | <50 | | | | | | | | | | |

**Supplemental Table S7: Predictive utility of LLM-inferred PHQ-9 scores for antidepressant switching and future psychiatric ED visits.** Andersen–Gill Cox proportional hazards models estimating the risk of future antidepressant switching or augmentation and psychiatric emergency department (ED) visits. Predictors include patient-reported PHQ-9, clinician-assessed SRA, and LLM-inferred PHQ-9. Models were conducted separately in two datasets: notes containing a patient-reported PHQ-9 and notes containing a clinician-assessed SRA. Predictors were also evaluated in joint models that included the alternative measure (e.g., Obs. PHQ-9 + LLM PHQ-9 or Obs. SRA + LLM PHQ-9), as indicated in the "**Joint w/**" column. To address potential violations of the independence assumption for recurrent events, additional analyses restricted outcomes to the first event per patient; these results are indicated by "*First*" in the "**Elig. Event**" column. All models were clustered by patient to account for repeated measures and outcomes. Statistical significance was defined using a Bonferroni-corrected threshold (p<0.05/8 *). Model discrimination was assessed using the concordance index (C-index).

| | | | Anti-depressant Switching | | | Psych. ED Visit | | |
|---|---|---|---|---|---|---|---|---|
| **Score** | **Joint w/** | **Elig. Event** | **exp(β)** | **pval** | **C-Index** | **exp(β)** | **pval** | **C-Index** |
| *Obs. PHQ-9 vs. LLM PHQ-9* | | | | | | | | |
| ***Obs. PHQ-9*** | ***NA*** | *All* | 1.06<br>(1.04-1.07) | 2.37e-12* | 0.60<br>(0.58-0.63) | 1.07<br>(1.04-1.11) | 1.44e-6* | 0.65<br>(0.60-0.71) |
| | | *First* | 1.05<br>(1.04-1.06) | 3.14e-18* | 0.60<br>(0.58-0.62) | 1.09<br>(1.05-1.12) | 3.14e-8* | 0.67<br>(0.61-0.72) |
| | ***LLM PHQ-9*** | *All* | 1.03<br>(1.01-1.06) | 4.58e-3* | 0.61<br>(0.59-0.64) | 1.05<br>(1.00-1.10) | 3.19e-2 | 0.66<br>(0.60-0.71) |
| | | *First* | 1.03<br>(1.01-1.05) | 3.04e-4* | 0.61<br>(0.59-0.63) | 1.06<br>(1.02-1.11) | 8.33e-3 | 0.67<br>(0.62-0.72) |
| ***LLM PHQ-9*** | ***NA*** | *All* | 1.06<br>(1.05-1.08) | 6.08e-19* | 0.61<br>(0.58-0.63) | 1.08<br>(1.04-1.11) | 7.61e-6* | 0.63<br>(0.57-0.69) |
| | | *First* | 1.06<br>(1.05-1.07) | 3.07e-19* | 0.60<br>(0.58-0.62) | 1.08<br>(1.05-1.12) | 1.34e-6* | 0.64<br>(0.58-0.69) |
| | ***Obs. PHQ-9*** | *All* | 1.04<br>(1.01-1.06) | 7.90e-4* | 0.61<br>(0.59-0.64) | 1.04<br>(0.99-1.09) | 1.29e-01 | 0.66<br>(0.60-0.71) |
| | | *First* | 1.03<br>(1.02-1.05) | 2.14e-4* | 0.61<br>(0.59-0.63) | 1.03<br>(0.98-1.09) | 1.97e-01 | 0.67<br>(0.62-0.72) |
| *Obs. SRA vs. LLM PHQ-9* | | | | | | | | |
| ***Obs. SRA*** | ***NA*** | *All* | 1.57<br>(1.24-1.98) | 1.58e-4* | 0.56<br>(0.52-0.59) | 2.09<br>(1.12-3.87) | 1.96e-02 | 0.61<br>(0.51-0.71) |
| | | *First* | 1.46<br>(1.16-1.83) | 1.19e-3* | 0.54<br>(0.51-0.57) | 1.75<br>(0.96-3.18) | 6.64e-02 | 0.59<br>(0.49-0.68) |
| | ***LLM PHQ-9*** | *All* | 1.19<br>(0.94-1.52) | 1.47e-1 | 0.61<br>(0.57-0.65) | 1.52<br>(0.74-3.13) | 2.56e-01 | 0.64<br>(0.55-0.74) |
| | | *First* | 1.13<br>(0.89-1.43) | 3.35e-1 | 0.61<br>(0.57-0.64) | 1.17<br>(0.69-1.99) | 5.55e-01 | 0.65<br>(0.55-0.75) |
| ***LLM PHQ-9*** | ***NA*** | *All* | 1.08<br>(1.06-1.11) | 1.98e-12* | 0.61<br>(0.57-0.64) | 1.10<br>(1.03-1.17) | 3.69e-03* | 0.64<br>(0.53-0.74) |
| | | *First* | 1.08<br>(1.06-1.11) | 3.40e-12* | 0.60<br>(0.57-0.64) | 1.11<br>(1.04-1.18) | 1.98e-03* | 0.65<br>(0.55-0.75) |
| | ***Obs. SRA*** | *All* | 1.07<br>(1.05-1.10) | 3.37e-9* | 0.61<br>(0.57-0.65) | 1.08<br>(1.00-1.16) | 5.74e-02 | 0.64<br>(0.55-0.74) |
| | | *First* | 1.08<br>(1.05-1.10) | 2.50e-9* | 0.61<br>(0.57-0.64) | 1.10<br>(1.03-1.17) | 3.91e-03* | 0.65<br>(0.55-0.75) |

**Supplemental Table S8: Predictive utility of LLM-inferred HAM-D scores for antidepressant switching and future psychiatric ED visits.** Andersen–Gill Cox proportional hazards models estimating the risk of future antidepressant switching or augmentation and psychiatry-associated emergency department (ED) visits. Predictors include patient-reported PHQ-9, clinician-assessed SRA, and LLM-inferred HAM-D. Models were conducted separately in two datasets: notes containing a patient-reported PHQ-9 and notes containing a clinician-assessed SRA. Predictors were also evaluated in joint models that included the alternative measure (e.g., Obs. PHQ-9 + LLM HAM-D or observed SRA + LLM HAM-D), as indicated in the "**Joint w/**" column. To address potential violations of the independence assumption for recurrent events, additional analyses restricted outcomes to the first event per patient; these results are indicated by "*First*" in the "**Elig. Event**" column. All models were clustered by patient to account for repeated measures and outcomes. Statistical significance was defined using a Bonferroni-corrected threshold (p<0.05/8 *). Model discrimination was assessed using the concordance index (C-index).

| Score | Joint w/ | Elig. Event | Anti-depressant Switching exp(β) | Anti-depressant Switching pval | Anti-depressant Switching C-Index | Psych. ED Visit exp(β) | Psych. ED Visit pval | Psych. ED Visit C-Index |
|---|---|---|---|---|---|---|---|---|
| *Obs. PHQ-9 vs. LLM HAM-D* | | | | | | | | |
| ***Obs. PHQ-9*** | ***NA*** | *All* | 1.06 (1.04-1.07) | 2.37e-12* | 0.60 (0.58-0.63) | 1.07 (1.04-1.11) | 1.44e-6* | 0.65 (0.60-0.71) |
| | | *First* | 1.05 (1.04-1.06) | 3.14e-18* | 0.60 (0.58-0.62) | 1.09 (1.05-1.12) | 3.14e-8* | 0.67 (0.61-0.72) |
| | ***LLM HAM-D*** | *All* | 1.04 (1.01-1.06) | 1.67e-3* | 0.61 (0.59-0.64) | 1.04 (1.00-1.08) | 8.14e-2 | 0.67 (0.61-0.72) |
| | | *First* | 1.03 (1.01-1.05) | 2.16e-4* | 0.61 (0.59-0.63) | 1.05 (1.00-1.10) | 3.26e-2 | 0.68 (0.63-0.73) |
| ***LLM HAM-D*** | ***NA*** | *All* | 1.06 (1.04-1.07) | 3.12e-18* | 0.60 (0.58-0.62) | 1.08 (1.05-1.11) | 3.17e-7* | 0.65 (0.60-0.71) |
| | | *First* | 1.05 (1.04-1.07) | 1.62e-19* | 0.60 (0.58-0.62) | 1.09 (1.06-1.12) | 1.36e-8* | 0.66 (0.61-0.72) |
| | ***Obs. PHQ-9*** | *All* | 1.03 (1.01-1.05) | 1.30e-3* | 0.61 (0.59-0.64) | 1.05 (1.01-1.10) | 1.40e-2 | 0.67 (0.61-0.72) |
| | | *First* | 1.03 (1.02-1.05) | 5.96e-5* | 0.61 (0.59-0.63) | 1.05 (1.01-1.10) | 1.84e-2 | 0.68 (0.63-0.73) |
| *Obs. SRA vs. LLM HAM-D* | | | | | | | | |
| ***Obs. SRA*** | ***NA*** | *All* | 1.57 (1.24-1.98) | 1.58e-4* | 0.56 (0.52-0.59) | 2.09 (1.12-3.87) | 1.96e-2 | 0.61 (0.51-0.71) |
| | | *First* | 1.46 (1.16-1.83) | 1.19e-3* | 0.54 (0.51-0.57) | 1.75 (0.96-3.18) | 6.64e-2 | 0.59 (0.49-0.68) |
| | ***LLM HAM-D*** | *All* | 1.26 (1.00-1.59) | 5.03e-2 | 0.60 (0.57-0.64) | 1.40 (0.73-2.68) | 3.11e-1 | 0.68 (0.60-0.77) |
| | | *First* | 1.20 (0.95-1.52) | 1.23e-1 | 0.60 (0.56-0.63) | 1.12 (0.65-1.92) | 6.80e-1 | 0.68 (0.59-0.78) |
| ***LLM HAM-D*** | ***NA*** | *All* | 1.07 (1.04-1.09) | 3.63e-9* | 0.60 (0.56-0.64) | 1.10 (1.05-1.16) | 9.02e-5* | 0.68 (0.59-0.76) |
| | | *First* | 1.06 (1.04-1.08) | 2.17e-8* | 0.59 (0.56-0.63) | 1.11 (1.05-1.17) | 2.18e-4* | 0.68 (0.59-0.78) |
| | ***Obs. SRA*** | *All* | 1.06 (1.03-1.08) | 4.56e-7* | 0.60 (0.57-0.64) | 1.09 (1.03-1.15) | 2.50e-3* | 0.68 (0.60-0.77) |
| | | *First* | 1.06 (1.03-1.08) | 1.39e-6* | 0.60 (0.56-0.63) | 1.10 (1.04-1.16) | 3.91e-4* | 0.68 (0.59-0.78) |

**Supplemental Table S9: Predictive utility of LLM-inferred CGI-S scores for antidepressant switching and future psychiatric ED visits.** Andersen–Gill Cox proportional hazards models estimating the risk of future antidepressant switching or augmentation and psychiatry-associated emergency department (ED) visits. Predictors include patient-reported PHQ-9, clinician-assessed SRA, and LLM-inferred CGI-S. Models were conducted separately in two datasets: notes containing a patient-reported PHQ-9 and notes containing a clinician-assessed SRA. Predictors were also evaluated in joint models that included the alternative measure (e.g., Obs. PHQ-9 + LLM CGI-S or observed SRA + LLM CGI-S), as indicated in the "**Joint w/**" column. To address potential violations of the independence assumption for recurrent events, additional analyses restricted outcomes to the first event per patient; these results are indicated by "*First*" in the "**Elig. Event**" column. All models were clustered by patient to account for repeated measures and outcomes. Statistical significance was defined using a Bonferroni-corrected threshold (p<0.05/8 *). Model discrimination was assessed using the concordance index (C-index).

| Score | Joint w/ | Elig. Event | Anti-depressant Switching exp(β) | pval | C-Index | Psych. ED Visit exp(β) | pval | C-Index |
|---|---|---|---|---|---|---|---|---|
| *Obs. PHQ-9 vs. LLM CGI-S* | | | | | | | | |
| ***Obs. PHQ-9*** | ***NA*** | *All* | 1.06 (1.04-1.07) | 2.37e-12* | 0.60 (0.58-0.63) | 1.07 (1.04-1.11) | 1.44e-6* | 0.65 (0.60-0.71) |
| | | *First* | 1.05 (1.04-1.06) | 3.14e-18* | 0.60 (0.58-0.62) | 1.09 (1.05-1.12) | 3.14e-8* | 0.67 (0.61-0.72) |
| | ***LLM CGI-S*** | *All* | 1.03 (1.01-1.06) | 9.59e-4* | 0.61 (0.59-0.64) | 1.05 (1.01-1.09) | 9.04e-3 | 0.66 (0.60-0.71) |
| | | *First* | 1.03 (1.02-1.05) | 3.62e-5* | 0.61 (0.59-0.63) | 1.07 (1.03-1.11) | 1.34e-3* | 0.67 (0.62-0.72) |
| ***LLM CGI-S*** | ***NA*** | *All* | 1.35 (1.26-1.45) | 3.69e-17* | 0.60 (0.58-0.62) | 1.45 (1.21-1.74) | 4.49e-5* | 0.63 (0.57-0.68) |
| | | *First* | 1.32 (1.24-1.40) | 7.71e-18* | 0.59 (0.57-0.61) | 1.48 (1.24-1.77) | 1.47e-5* | 0.63 (0.57-0.68) |
| | ***Obs. PHQ-9*** | *All* | 1.20 (1.09-1.31) | 1.01e-4* | 0.61 (0.59-0.64) | 1.21 (0.96-1.51) | 1.01e-1 | 0.66 (0.60-0.71) |
| | | *First* | 1.18 (1.09-1.28) | 7.78e-5* | 0.61 (0.59-0.63) | 1.17 (0.93-1.48) | 1.69e-1 | 0.67 (0.62-0.72) |
| *Obs. SRA vs. LLM CGI-S* | | | | | | | | |
| ***Obs. SRA*** | ***NA*** | *All* | 1.57 (1.24-1.98) | 1.58e-4* | 0.56 (0.52-0.59) | 2.09 (1.12-3.87) | 1.96e-2 | 0.61 (0.51-0.71) |
| | | *First* | 1.46 (1.16-1.83) | 1.19e-3* | 0.54 (0.51-0.57) | 1.75 (0.96-3.18) | 6.64e-2 | 0.59 (0.49-0.68) |
| | ***LLM CGI-S*** | *All* | 1.27 (1.02-1.59) | 3.41e-2 | 0.61 (0.57-0.65) | 1.59 (0.82-3.08) | 1.70e-1 | 0.65 (0.56-0.75) |
| | | *First* | 1.20 (0.96-1.51) | 1.10e-1 | 0.61 (0.57-0.64) | 1.31 (0.76-2.28) | 3.36e-1 | 0.65 (0.56-0.75) |
| ***LLM CGI-S*** | ***NA*** | *All* | 1.44 (1.28-1.62) | 1.15e-9* | 0.61 (0.57-0.65) | 1.54 (1.18-2.02) | 1.43e-3* | 0.65 (0.56-0.73) |
| | | *First* | 1.43 (1.28-1.59) | 2.31e-10* | 0.60 (0.57-0.64) | 1.54 (1.17-2.03) | 2.18e-3* | 0.65 (0.56-0.74) |
| | ***Obs. SRA*** | *All* | 1.39 (1.24-1.56) | 3.45e-8* | 0.61 (0.57-0.65) | 1.42 (1.05-1.90) | 2.09e-2 | 0.65 (0.56-0.75) |
| | | *First* | 1.39 (1.24-1.56) | 8.04e-9* | 0.61 (0.57-0.64) | 1.47 (1.13-1.91) | 4.25e-3* | 0.65 (0.56-0.75) |

**Table S10. Consistency of score conversions between LLM-inferred and observed psychometric measures across clinics.** Score conversion weights were estimated for LLM-inferred PHQ-9, HAM-D, and CGI-S scores relative to patient-reported PHQ-9 scores and the mean study-clinician chart review CGI-S. Weights were derived using mixed-effects linear models in a meta-analytic framework, with clinic-level random effects (random intercepts and slopes) and subject-level random effects. Reported estimates include the conversion weights with 95% confidence intervals, the standard deviation of true between-clinic variation (τ), and the proportion of variance attributable to between-clinic heterogeneity ($I^2$).

| | ***Outcome*** | | | | | |
|---|---|---|---|---|---|---|
| | ***Obs. PHQ-9 (n=29 clinics)*** | | | ***Avg. R CGI-S (n=12 clinics)*** | | |
| **Input** | **weight** | **$\tau^2$** | **$I^2$** | **weight** | **$\tau^2$** | **$I^2$** |
| **LLM PHQ-9** | 0.64 (0.58-0.70) | 0.08 | 0.00 | 0.16 (0.12-0.20) | 0.05 | 0.01 |
| **LLM HAM-D** | 0.57 (0.53-0.61) | 0.04 | 0.00 | 0.16 (0.14-0.19) | 0.02 | 0.00 |
| **LLM CGI-S** | 2.77 (2.56-2.98) | 0.21 | 0.00 | 0.72 (0.59-0.85) | 0.07 | 0.02 |

**Supplemental Table S11. Convergent and predictive validity of LLM-inferred depression severity comparing GPT-5-mini and GPT-5.2.** This table reports point estimates and 95% confidence intervals for metrics evaluating LLM-inferred PHQ-9, HAM-D, and CGI-S produced by GPT-5-mini and GPT-5.2. For patient-reported PHQ-9 (Obs. PHQ-9), we report agreement with PHQ-9 ($\kappa_{phq9}$; i.e. Cohen's κ with quadratic weighting), $r_{overall}$ and $r_{repeated}$ correlations, and AUC for Obs. PHQ-9≥10. For clinician-rated CGI-S, we report agreement between LLM-inferred and each of the two study-clinician CGI-S ratings ($\kappa_{R1}$ and $\kappa_{R2}$), as well as overall ($r_{overall}$) and repeated-measures ($r_{repeated}$) correlations with mean study-clinician CGI-S ratings. Predictive validity is evaluated using Andersen-Gill Cox proportional hazards models for antidepressant switching/augmentation (AD switching) and psychiatric emergency department visits (psych. ED visits), reporting hazard ratios, p-values, and C-indices. Analysis was done on all notes with an associated patient-reported PHQ-9 (n=3,757 notes, 1,480 patients).

| Exp. | Metric | LLM PHQ-9 GPT-5-mini | LLM PHQ-9 GPT-5.2 | LLM HAM-D GPT-5-mini | LLM HAM-D GPT-5.2 | LLM CGI-S GPT-5-mini | LLM CGI-S GPT-5.2 |
|---|---|---|---|---|---|---|---|
| *Convergent Validity* | | | | | | | |
| ***Obs. PHQ-9*** | $\kappa_{phq9}$ | **0.64 (0.62-0.66)** | **0.64 (0.62-0.66)** | NA | NA | NA | NA |
| | $r_{overall}$ | 0.66 (0.64-0.68) | **0.67 (0.65-0.68)** | 0.57 (0.55-0.59) | 0.65 (0.64-0.67) | 0.60 (0.58-0.62) | 0.63 (0.61-0.65) |
| | $r_{repeated}$ | 0.47 (0.44-0.51) | **0.50 (0.47-0.53)** | 0.41 (0.37-0.44) | 0.48 (0.45-0.51) | 0.39 (0.35-0.42) | 0.45 (0.42-0.48) |
| | *AUC* | **0.83 (0.81-0.84)** | **0.83 (0.81-0.84)** | 0.79 (0.77-0.80) | 0.82 (0.81-0.84) | 0.79 (0.78-0.81) | 0.80 (0.79-0.82) |
| ***Rater CGI-S*** | $\kappa_{rater1}$ | NA | NA | NA | NA | 0.74 (0.64-0.81) | **0.79 (0.70-0.85)** |
| | $\kappa_{rater2}$ | NA | NA | NA | NA | **0.70 (0.59-0.77)** | 0.67 (0.58-0.77) |
| ***Avg. Rater CGI-S*** | $r_{overall}$ | 0.83 (0.76-0.88) | **0.88 (0.83-0.91)** | 0.78 (0.71-0.84) | **0.88 (0.84-0.92)** | 0.85 (0.79-0.89) | 0.86 (0.80-0.90) |
| | $r_{repeated}$ | 0.69 (0.57-0.78) | 0.78 (0.69-0.85) | 0.67 (0.55-0.77) | **0.82 (0.74-0.88)** | 0.73 (0.62-0.81) | 0.79 (0.70-0.85) |
| *Predictive Validity (vs. Obs. PHQ-9)* | | | | | | | |
| ***AD Switching*** | $exp(\beta)$ | 1.06 (1.05-1.08) | 1.06 (1.05-1.08) | 1.06 (1.04-1.07) | 1.06 (1.04-1.07) | 1.32 (1.23-1.42) | 1.35 (1.26-1.45) |
| | *pval* | 8.59e-18 | 6.08e-19 | 3.35e-16 | 3.12e-18 | 2.20e-14 | 3.69e-17 |
| | *C-index* | 0.60 (0.58-0.62) | **0.61 (0.58-0.63)** | 0.60 (0.57-0.62) | 0.60 (0.58-0.62) | 0.59 (0.57-0.61) | 0.60 (0.58-0.62) |
| ***Psych. ED Visits*** | $exp(\beta)$ | 1.09 (1.05-1.13) | 1.08 (1.04-1.11) | 1.10 (1.06-1.14) | 1.08 (1.05-1.11) | 1.44 (1.20-1.74) | 1.45 (1.21-1.74) |
| | *pval* | 3.25e-6 | 7.61e-6 | 1.61e-7 | 3.17e-7 | 1.03e-4 | 4.49e-5 |
| | *C-index* | 0.64 (0.58-0.69) | 0.63 (0.57-0.69) | **0.66 (0.60-0.72)** | 0.65 (0.60-0.71) | 0.62 (0.57-0.67) | 0.63 (0.57-0.68) |

### Supplemental Figures:

**Figure S1: Eligibility criteria for clinical notes and the available observed psychometric. A)** Overview of the clinical note selection process used to construct two datasets: the MDD Dataset and the Diagnosis-Stratified Cohort Dataset. The schematic begins with the initial pool of retrieved notes (see Note Retrieval and Eligibility subsection in Supplement) and outlines the eligibility criteria applied to define each dataset. **B)** The number of clinical notes (top) and unique patients (bottom) in the *MDD Note Dataset* with available psychometric scores, including patient-reported PHQ-9 completed within the 7 days prior to the visit, treating clinician-assessed suicide risk assessments (SRA), and study-clinician chart review assessments (CGI-S). **C)** Illustration of overlap among notes containing observed PHQ-9, SRA, and CGI-S (i.e. study-clinician chart review) within the *MDD Note dataset*.

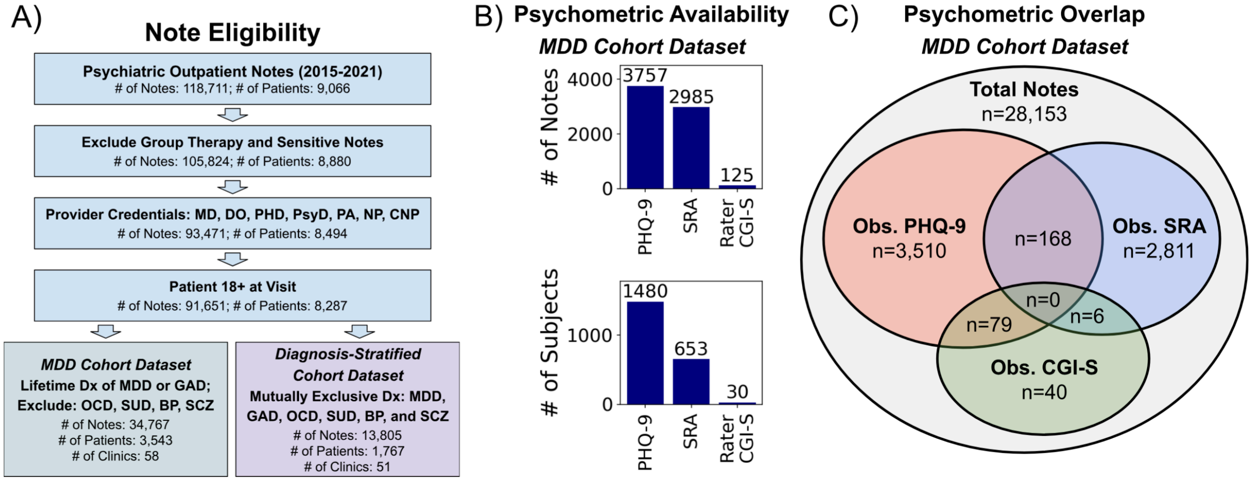

**Figure S2. Locally Weighted Scatterplot Smoothing (LOWESS) curves comparing LLM-inferred and observed psychometric scores.** Each panel displays a LOWESS fit with 95% confidence intervals derived from bootstrap resampling (n=1,000), with individual patients shown as scatter points. **A)** LOWESS fit of LLM-inferred versus patient-reported PHQ-9 scores, where the dashed line represents perfect agreement (identity line). **B)** Absolute error (|LLM-inferred − Observed|) across the observed PHQ-9 range. **C)** LOWESS fit of LLM-inferred versus study-clinician CGI-S ratings, with the dashed identity line indicating perfect agreement. **D)** Absolute error (|LLM-inferred − Observed|) across the observed CGI-S range.

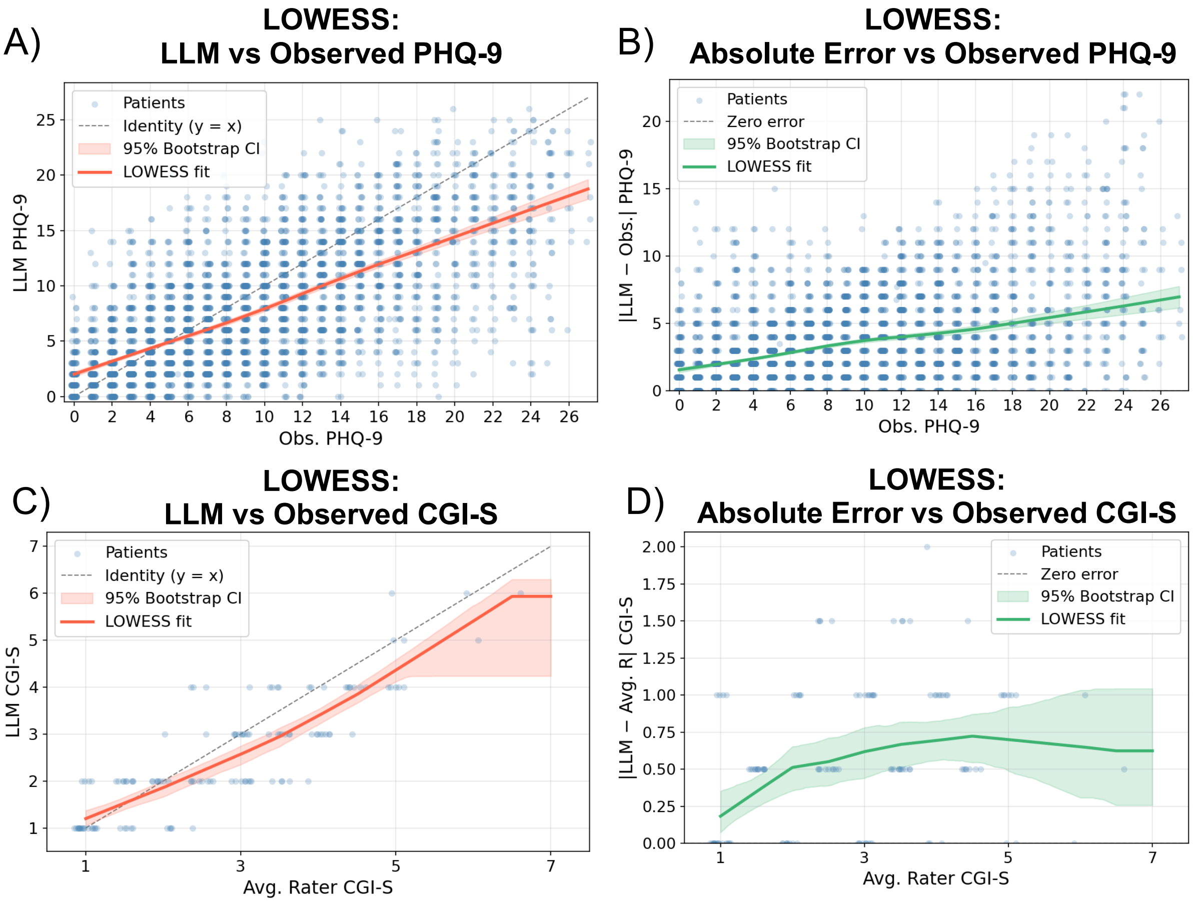

**Supplemental Figure S3: LLM sensitivity to blinded SRA section.** Receiver operating characteristic curves are shown for clinician-assessed suicide risk (SRA≥2; i.e. greater-than-minimal suicide risk) at each visit, based on LLM-inferred depression severity scores (e.g., LLM PHQ-9, LLM HAM-D, LLM CGI-S), with and without blinding of the SRA section (see SRA Blinding Subsection in Supplement for details on how the SRA section was redacted). Analyses were conducted on all psychiatric clinical notes with an assigned SRA score in the *MDD dataset* (n = 2,985 notes, 653 patients).

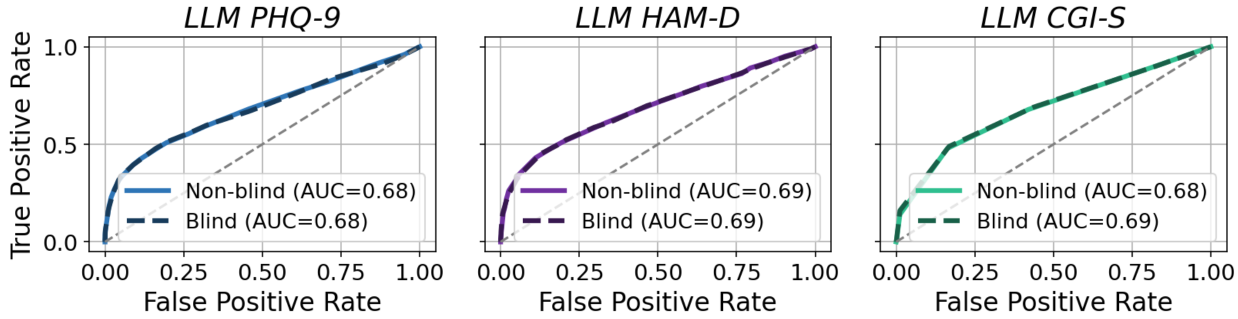


**Supplemental Figure S4: Survival curves for antidepressant switching or augmentation using observed and LLM-inferred depression severity measures** Case-control Kaplan-Meier survival curves for time to antidepressant switching or augmentation were generated to compare risk stratification using: (1) patient-reported PHQ-9 scores (cases ≥20; controls <10) versus LLM PHQ-9 scores (cases ≥17; controls <8) or LLM HAM-D (cases ≥20; controls <12), and (2) clinician-rated suicide risk assessment (cases ≥2; controls <2) versus LLM PHQ-9 scores (cases ≥8; controls <8) or (LLM HAM-D ≥13; controls <13). Shaded regions denote 95% confidence intervals for observed PHQ-9 and suicide risk assessment case-control curves. Corresponding percentage cutoffs for each definition are reported in Supplemental Table S5.

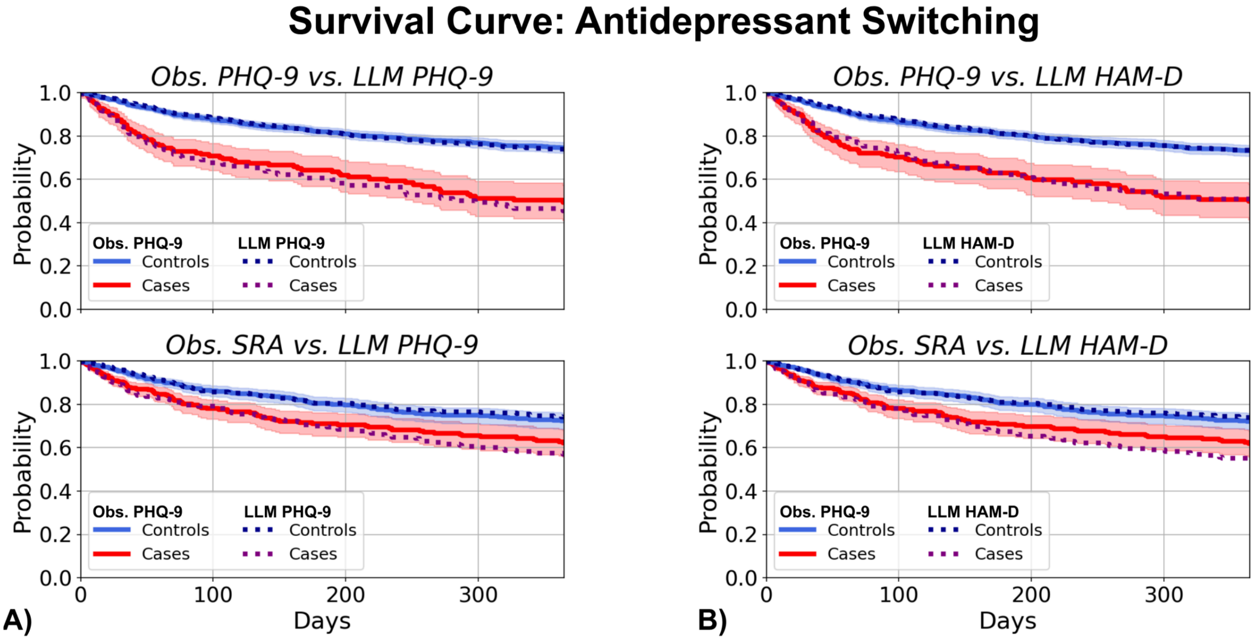

**Supplemental Figure S5: Convergent validity of LLM-inferred depression severity scores across prompt types and GPT versions. A)** Overall and repeated-measures correlations between LLM-inferred depression severity scores (PHQ-9, HAM-D, and CGI-S) generated using different prompt types (name-only; full-scale [default]; item-level; and CGI-S short) and patient-reported PHQ-9 and mean study-clinician CGI-S ratings. **B)** Overall and repeated-measures correlations between LLM-inferred depression severity scores across GPT versions (gpt-4o-mini, gpt-4o, gpt-5-mini, gpt-5, and gpt-5.2) and to the same psychometrics as denoted above. Sensitivity analyses were conducted using clinical notes in the *MDD dataset* with either 1) PHQ-9 and SRA scores or 2) chart reviewer ratings (n=293 notes, 148 patients).

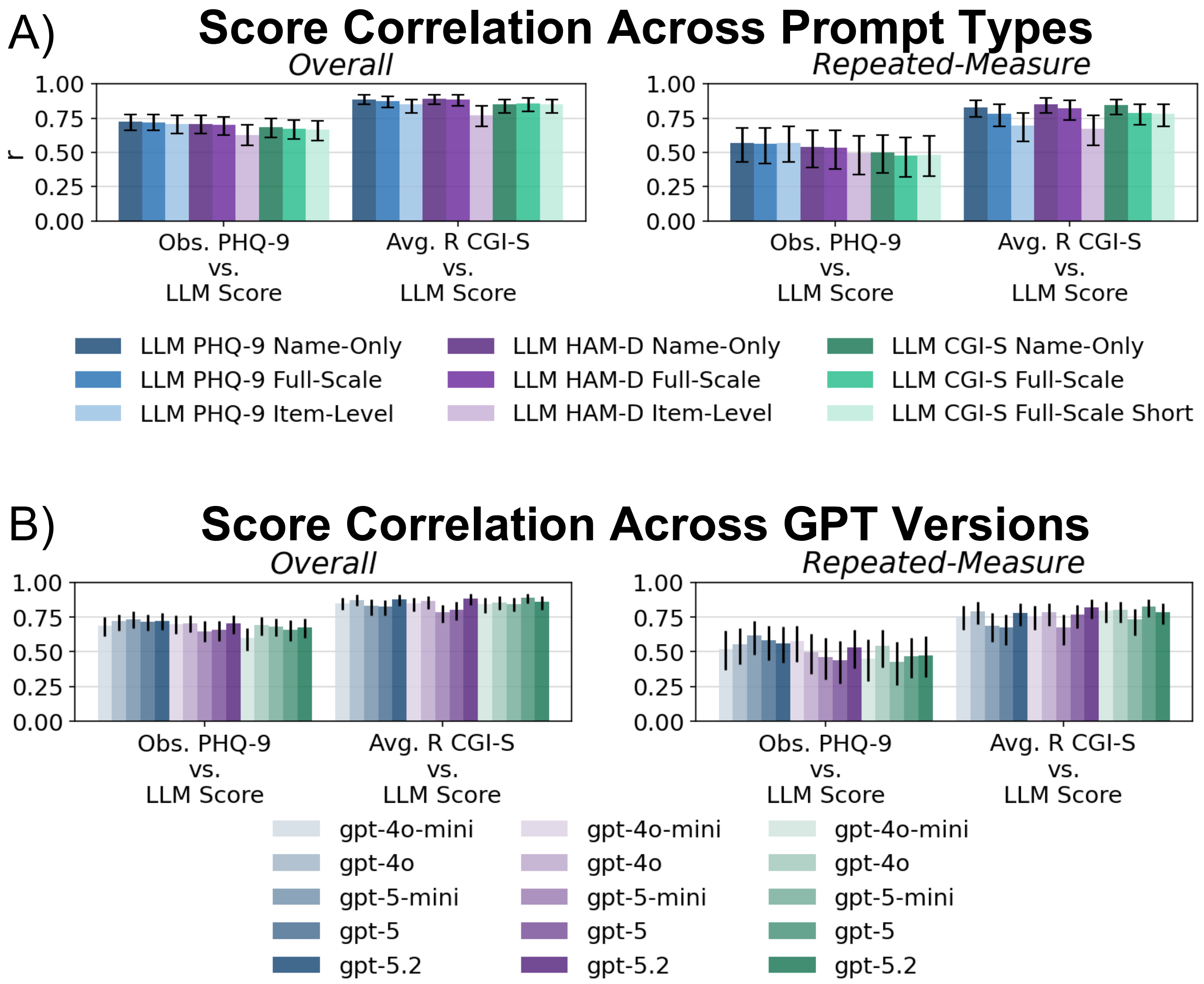

**Supplemental Figure S6: LLM score distribution and correlation across score and prompt type.** Illustrations of **A)** the observed distributions and **B)** overall Pearson correlations of LLM-inferred depression scores (PHQ-9, HAM-D, and CGI-S) across three prompt variants—Name-Only, Full-Scale, and Item-Level—including a shortened CGI-S Full-Scale prompt (Full-Scale Short). Sensitivity analyses were conducted using clinical notes in the *MDD dataset* with either 1) PHQ-9 and SRA scores or 2) chart reviewer ratings (n=293 notes, 148 patients).

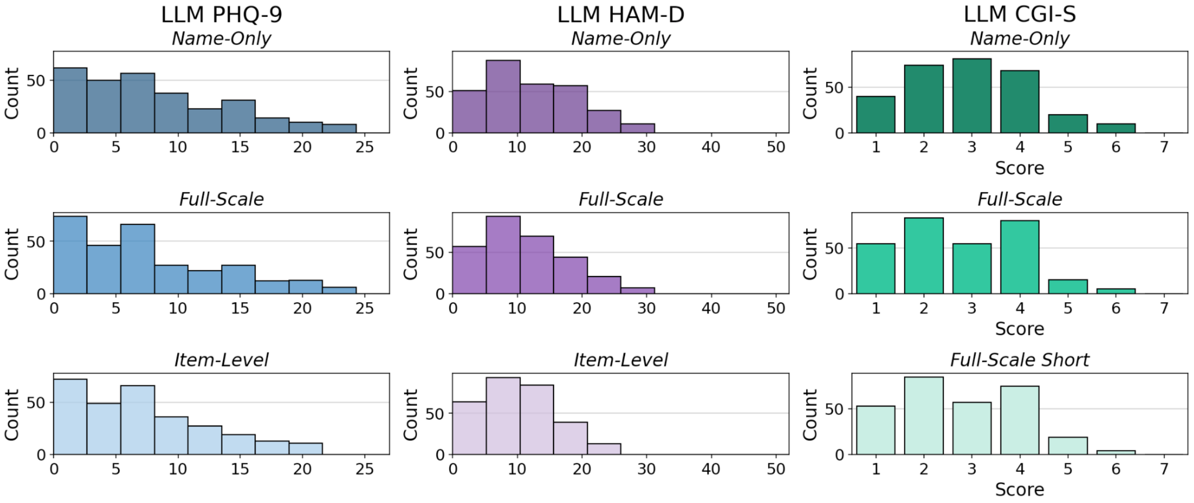


**A)**

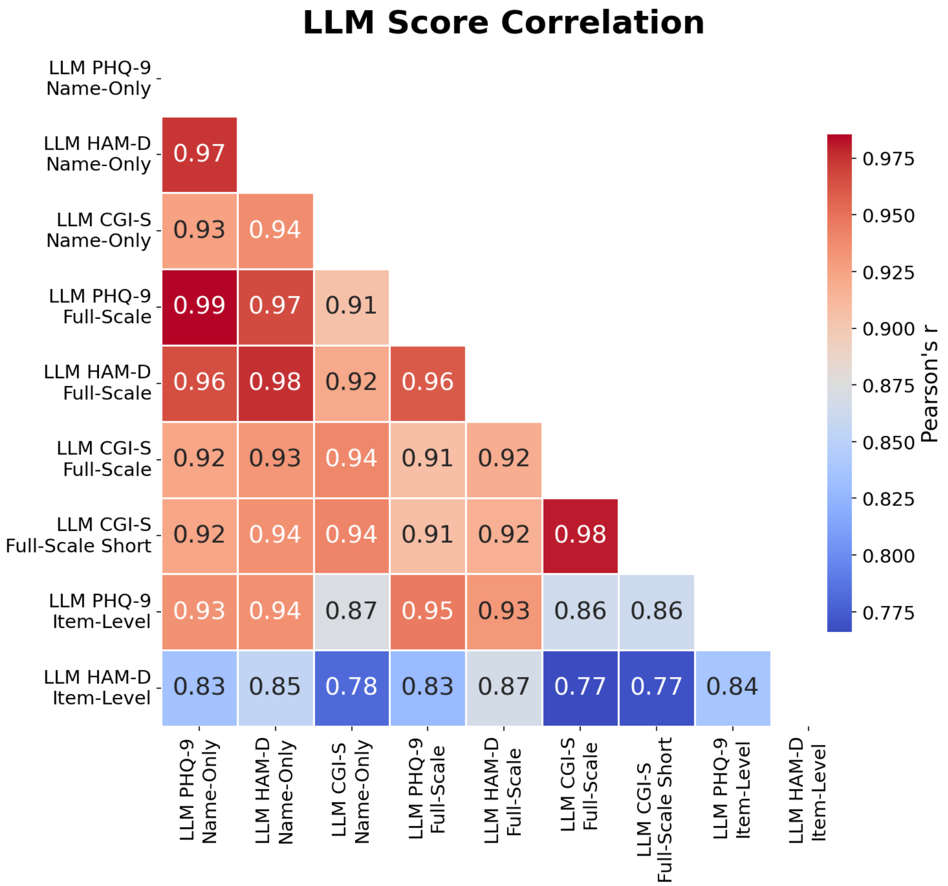


**B)**
