## Supplementary material for "Multi-Criteria Validation of LLM-Inferred Depression Severity from Outpatient Psychiatry Notes": LLM Prompting

### *Full-Scale* LLM 9-Item Patient Health Questionnaire Scale Prompt

**System Prompt**:
“You are an expert psychiatrist analyzing clinical notes”

**User Prompt**:
“Based on the provided clinical note, estimate the patient's total Patient Health Questionnaire-9 (PHQ-9) score (range: 0-27) at the current visit.

[SCALE_TEXT]

Clinical Note from Current Visit:
[CLINICAL_NOTE]”

**Output Structure**:

```
{
  "name": "PHQ9",
  "schema": {
    "type": "object",
    "properties": {
      "PHQ-9_total": {
        "type": "integer",
        "description": "Patient Health Questionnaire-9 (PHQ-9) total score ranging from 0 to 27, where 0–4 = minimal depression, 5–9 = mild, 10–14 = moderate, 15–19 = moderately severe, and 20–27 = severe depression",
        "minimum": 0,
        "maximum": 27
      }
    },
    "required": ["PHQ-9_total"],
    "additionalProperties": false
  }
}
```

#### *Name-Only* LLM 9-Item Patient Health Questionnaire Scale Prompt

**System Prompt**:
“You are an expert psychiatrist analyzing clinical notes”

**User Prompt**:
“Based on the provided clinical note, estimate the patient's total Patient Health Questionnaire-9 (PHQ-9) score (range: 0-27) at the current visit.

Clinical Note from Current Visit:
[CLINICAL_NOTE]”

**Output Structure**:

```
{
  "name": "PHQ9",
  "schema": {
    "type": "object",
    "properties": {
      "PHQ-9_total": {
        "type": "integer",
        "description": "Patient Health Questionnaire-9 (PHQ-9) total score ranging from 0 to 27, where 0–4 = minimal depression, 5–9 = mild, 10–14 = moderate, 15–19 = moderately severe, and 20–27 = severe depression",
        "minimum": 0,
        "maximum": 27
      }
    },
    "required": ["PHQ-9_total"],
    "additionalProperties": false
  }
}
```

#### *Item-Level* LLM 9-Item Patient Health Questionnaire Scale Prompt

**System Prompt**:
“You are an expert psychiatrist analyzing clinical notes”

**User Prompt**:
“Based on the provided clinical note, estimate the patient's itemized Patient Health Questionnaire-9 (PHQ-9) scores at the current visit.

[SCALE_TEXT]

Clinical Note from Current Visit:
[CLINICAL_NOTE]”

**Output Structure**:


```
{
 "name": "PHQ-9",
 "schema": {
  "type": "object",
  "properties": {
   "PHQ-9_items": {
    "type": "object",
    "properties": {
     "anhedonia": { "type": "integer", "minimum": 0, "maximum": 3 },
     "depressed_mood": { "type": "integer", "minimum": 0, "maximum": 3 },
     "sleep_disturbance": { "type": "integer", "minimum": 0, "maximum": 3 },
     "fatigue": { "type": "integer", "minimum": 0, "maximum": 3 },
     "appetite_change": { "type": "integer", "minimum": 0, "maximum": 3 },
     "worthlessness_or_guilt": { "type": "integer", "minimum": 0, "maximum": 3 },
     "concentration_difficulty": { "type": "integer", "minimum": 0, "maximum": 3 },
     "psychomotor_change": { "type": "integer", "minimum": 0, "maximum": 3 },
     "suicidal_ideation": { "type": "integer", "minimum": 0, "maximum": 3 }
    },
    "required": [
     "anhedonia",
     "depressed_mood",
     "sleep_disturbance",
     "fatigue",
     "appetite_change",
     "worthlessness_or_guilt",
     "concentration_difficulty",
     "psychomotor_change",
     "suicidal_ideation"
    ],
```

```
        "additionalProperties": false
      }
    },
    "required": ["PHQ-9_items"],
    "additionalProperties": false
  }
}
```

### *Full-Scale* LLM Hamilton Depression Rating Scale Prompt

**System Prompt**:
“You are an expert psychiatrist analyzing clinical notes”

**User Prompt**:
“Based on the provided clinical note, estimate the patient's total Hamilton Depression Rating Scale (HAM-D) score (range: 0-52) at the current visit.

[SCALE_TEXT]

Clinical Note from Current Visit:
[CLINICAL_NOTE]”

**Output Structure**:
```
{
  "name": "HAM-D",
  "schema": {
    "type": "object",
    "properties": {
      "HAM-D_total": {
        "type": "integer",
        "description": "Hamilton Depression Rating Scale (HAM-D) total score ranging from 0 to 52, where 0–7 = normal, 8–13 = mild, 14–18 = moderate, 19–22 = severe, and ≥23 = very severe depression",
        "minimum": 0,
        "maximum": 52
      }
    },
    "required": ["HAM-D_total"],
    "additionalProperties": false
  }
}
```

#### *Name-Only* LLM Hamilton Depression Rating Scale Prompt

**System Prompt**:
“You are an expert psychiatrist analyzing clinical notes”

**User Prompt**:
“Based on the provided clinical note, estimate the patient's total Hamilton Depression Rating Scale (HAM-D) score (range: 0-52) at the current visit.

Clinical Note from Current Visit:
[CLINICAL_NOTE]”

**Output Structure**:

```
{
  "name": "HAM-D",
  "schema": {
    "type": "object",
    "properties": {
      "HAM-D_total": {
        "type": "integer",
        "description": "Hamilton Depression Rating Scale (HAM-D) total score ranging from 0 to 52, where 0–7 = normal, 8–13 = mild, 14–18 = moderate, 19–22 = severe, and ≥23 = very severe depression",
        "minimum": 0,
        "maximum": 52
      }
    },
    "required": ["HAM-D_total"],
    "additionalProperties": false
  }
}
```

### *Item-Level* LLM Hamilton Depression Rating Scale Prompt

**System Prompt**:
"You are an expert psychiatrist analyzing clinical notes"

**User Prompt**:
"Based on the provided clinical note, estimate the patient's itemized Hamilton Depression Rating Scale (HAM-D) scores at the current visit.

[SCALE_TEXT]

Clinical Note from Current Visit:
[CLINICAL_NOTE]"

**Output Structure**:

```
{
 "name": "HAM-D",
 "schema": {
  "type": "object",
  "properties": {
   "HAM-D_items": {
    "type": "object",
    "properties": {
     "depressed_mood": { "type": "integer", "minimum": 0, "maximum": 4 },
     "feelings_of_guilt": { "type": "integer", "minimum": 0, "maximum": 4 },
     "suicide": { "type": "integer", "minimum": 0, "maximum": 4 },
     "insomnia_early": { "type": "integer", "minimum": 0, "maximum": 2 },
     "insomnia_middle": { "type": "integer", "minimum": 0, "maximum": 2 },
     "insomnia_late": { "type": "integer", "minimum": 0, "maximum": 2 },
     "work_and_activities": { "type": "integer", "minimum": 0, "maximum": 4 },
     "psychomotor_retardation": { "type": "integer", "minimum": 0, "maximum": 4 },
     "psychomotor_agitation": { "type": "integer", "minimum": 0, "maximum": 4 },
     "anxiety_psychic": { "type": "integer", "minimum": 0, "maximum": 4 },
     "anxiety_somatic": { "type": "integer", "minimum": 0, "maximum": 4 },
     "somatic_symptoms_gastrointestinal": { "type": "integer", "minimum": 0, "maximum": 2 },
     "somatic_symptoms_general": { "type": "integer", "minimum": 0, "maximum": 2 },
     "genital_symptoms": { "type": "integer", "minimum": 0, "maximum": 2 },
     "hypochondriasis": { "type": "integer", "minimum": 0, "maximum": 4 },
     "weight_loss": { "type": "integer", "minimum": 0, "maximum": 2 },
     "insight": { "type": "integer", "minimum": 0, "maximum": 2 }
    },
    "required": [
     "depressed_mood",
     "feelings_of_guilt",
```

"suicide",
"insomnia_early",
"insomnia_middle",
"insomnia_late",
"work_and_activities",
"psychomotor_retardation",
"psychomotor_agitation",
"anxiety_psychic",
"anxiety_somatic",
"somatic_symptoms_gastrointestinal",
"somatic_symptoms_general",
"genital_symptoms",
"hypochondriasis",
"weight_loss",
"insight"
],
"additionalProperties": false
}
},
"required": ["HAM-D_items"],
"additionalProperties": false
}
}

#### *Full-Scale (and Full-Scale Short)* LLM Clinical Global Impression – Severity Prompt

**System Prompt**:
“You are an expert psychiatrist analyzing clinical notes”

**User Prompt**:
“Based on the provided clinical note, estimate the patient's total Clinical Global Impression – Severity (CGI-S) score (range: 1-7) for Major Depressive Disorder at the current visit.

[SCALE_TEXT]

Clinical Note from Current Visit:
[CLINICAL_NOTE]”

**Output Structure**:

```
{
  "name": "CGI-S",
  "schema": {
    "type": "object",
    "properties": {
      "CGI-S_total": {
          "type": "integer",
          "minimum": 1,
          "maximum": 7,
          "description": "Clinical Global Impression - Severity Rating for Major Depressive Disorder ranging from 1-7, where 1 = normal, 2 = borderline ill, 3 = mildly ill, 4 = moderately ill, 5 = markedly ill, 6 = severely ill, 7 = among the most extremely ill patients."
      }
    },
    "required": ["CGI-S_total"],
    "additionalProperties": false
  }
}
```

### *Name-Only* LLM Clinical Global Impression – Severity Prompt

**System Prompt**:
“You are an expert psychiatrist analyzing clinical notes”

**User Prompt**:
“Based on the provided clinical note, estimate the patient's total Clinical Global Impression – Severity (CGI-S) score (range: 1-7) for Major Depressive Disorder at the current visit.

Clinical Note from Current Visit:
[CLINICAL_NOTE]”

**Output Structure**:

```
{
  "name": "CGI-S",
  "schema": {
    "type": "object",
    "properties": {
      "CGI-S_total": {
            "type": "integer",
            "minimum": 1,
            "maximum": 7,
            "description": "Clinical Global Impression - Severity Rating for Major Depressive Disorder ranging from 1-7, where 1 = normal, 2 = borderline ill, 3 = mildly ill, 4 = moderately ill, 5 = markedly ill, 6 = severely ill, 7 = among the most extremely ill patients."
      }
    },
    "required": ["CGI-S_total"],
    "additionalProperties": false
  }
}
```
