## Supplementary material for "Multi-Criteria Validation of LLM-Inferred Depression Severity from Outpatient Psychiatry Notes": Scale Text

### 9-Item Patient Health Questionnaire Scale Text

“Patient Health Questionnaire-9 (PHQ-9)

Over the last 2 weeks, how often have you been bothered by any of the following problems?

0 = Not at all
1 = Several days
2 = More than half the days
3 = Nearly every day

1. Little interest or pleasure in doing things
2. Feeling down, depressed, or hopeless
3. Trouble falling or staying asleep, or sleeping too much
4. Feeling tired or having little energy
5. Poor appetite or overeating
6. Feeling bad about yourself — or that you are a failure or have let yourself or your family down
7. Trouble concentrating on things, such as reading the newspaper or watching television
8. Moving or speaking so slowly that other people could have noticed? Or the opposite — being so fidgety or restless that you have been moving around a lot more than usual
9. Thoughts that you would be better off dead or of hurting yourself in some way”

*Source:*
Kroenke K, Spitzer RL, Williams JBW. The PHQ-9. *J Gen Intern Med*. 2001;16(9):606-613.

### Hamilton Depression Rating Scale Text

"Hamilton Depression Rating Scale (HDRS)

Instructions: for each item select the one "cue" which best characterizes the patient. Be sure to record the answers in the appropriate spaces (positions 0 through 4).

1. DEPRESSED MOOD (sadness, hopeless, helpless, worthless)
0 - Absent.
1 - These feeling states indicated only on questioning.
2 - These feeling states spontaneously reported verbally.
3 - Communicates feeling states non-verbally, i.e. through facial expression, posture, voice and tendency to weep.
4 - Patient reports virtually only these feeling states in his/her spontaneous verbal and non-verbal communication.

2. FEELINGS OF GUILT
0 - Absent.
1 - Self reproach, feels he/she has let people down.
2 - Ideas of guilt or rumination over past errors or sinful deeds.
3 - Present illness is a punishment. Delusions of guilt.
4 - Hears accusatory or denunciatory voices and/or experiences threatening visual hallucinations.

3. SUICIDE
0 - Absent.
1 - Feels life is not worth living.
2 - Wishes he/she were dead or any thoughts of possible death to self.
3 - Ideas or gestures of suicide.
4 - Attempts at suicide (any serious attempt rate 4).

4. INSOMNIA: EARLY IN THE NIGHT
0 - No difficulty falling asleep.
1 - Complains of occasional difficulty falling asleep, i.e. more than 1/2 hour.
2 - Complains of nightly difficulty falling asleep.

5. INSOMNIA: MIDDLE OF THE NIGHT
0 - No difficulty.
1 - Patient complains of being restless and disturbed during the night.
2 - Waking during the night – any getting out of bed rates 2 (except for purposes of voiding).

6. INSOMNIA: EARLY HOURS OF THE MORNING
0 - No difficulty.
1 - Waking in early hours of the morning but goes back to sleep.
2 - Unable to fall asleep again if he/she gets out of bed.

7. WORK AND ACTIVITIES
0 - No difficulty.
1 - Thoughts and feelings of incapacity, fatigue or weakness related to activities, work or hobbies.
2 - Loss of interest in activity, hobbies or work – either directly reported by the patient or indirect in listlessness, indecision and vacillation (feels he/she has to push self to work or activities).
3 - Decrease in actual time spent in activities or decrease in productivity. Rate 3 if the patient does not spend at least three hours a day in activities (job or hobbies) excluding routine chores.
4 - Stopped working because of present illness. Rate 4 if patient engages in no activities except routine chores, or if patient fails to perform routine chores unassisted.

8. RETARDATION (slowness of thought and speech, impaired ability to concentrate, decreased motor activity)
0 - Normal speech and thought.
1 - Slight retardation during the interview.
2 - Obvious retardation during the interview.
3 - Interview difficult.
4 - Complete stupor.

9. AGITATION
0 - None.
1 - Fidgetiness.
2 - Playing with hands, hair, etc.
3 - Moving about, can’t sit still.
4 - Hand wringing, nail biting, hair-pulling, biting of lips.

10. ANXIETY PSYCHIC
0 - No difficulty.
1 - Subjective tension and irritability.
2 - Worrying about minor matters.
3 - Apprehensive attitude apparent in face or speech.
4 - Fears expressed without questioning.

11. ANXIETY SOMATIC (physiological concomitants of anxiety) such as: gastro-intestinal – dry mouth, wind, indigestion, diarrhea, cramps, belching cardio-vascular – palpitations, headaches respiratory – hyperventilation, sighing urinary frequency sweating
0 - Absent.
1 - Mild.
2 - Moderate.
3 - Severe.
4 - Incapacitating.

12. SOMATIC SYMPTOMS GASTRO-INTESTINAL

0 - None.
1 - Loss of appetite but eating without staff encouragement. Heavy feelings in abdomen.
2 - Difficulty eating without staff urging. Requests or requires laxatives or medication for bowels or medication for gastro-intestinal symptoms.

13. GENERAL SOMATIC SYMPTOMS
0 - None.
1 - Heaviness in limbs, back or head. Backaches, headaches, muscle aches. Loss of energy and fatigability.
2 - Any clear-cut symptom rates 2.

14. GENITAL SYMPTOMS (symptoms such as loss of libido, menstrual disturbances)
0 - Absent.
1 - Mild.
2 - Severe.

15. HYPOCHONDRIASIS
0 - Not present.
1 - Self-absorption (bodily).
2 - Preoccupation with health.
3 - Frequent complaints, requests for help, etc.
4 - Hypochondriacal delusions.

16. LOSS OF WEIGHT (RATE EITHER a OR b)
a) According to the patient:
0 - No weight loss.
1 - Probable weight loss associated with present illness.
2 - Definite (according to patient) weight loss.
3 - Not assessed.

b) According to weekly measurements:
0 - Less than 1 lb weight loss in week.
1 - Greater than 1 lb weight loss in week.
2 - Greater than 2 lb weight los in week.
3 - Not assessed.

17 INSIGHT
0 - Acknowledges being depressed and ill.
1 - Acknowledges illness but attributes cause to bad food, climate, overwork, virus, need for rest, etc.
2 - Denies being ill at all."

*Source:*
Hamilton M. A rating scale for depression. J Neurol Neurosurg Psychiatry. 1960;23(1):56-62.

### Depression-Specific Clinical Global Impression – Severity Text

“Clinical Global Impression - Severity (Major Depressive Disorder)

Severity Ratings:
1. Normal - Depressive symptoms are rarely present and occur only in contextually appropriate circumstances; functioning is at or near full capacity.
2. Borderline ill - Depressive symptoms are few, intermittent, and mild; little or no interference with role functioning.
3. Mildly ill - Depressive symptoms are clearly present and distressing, but functioning is only minimally reduced.
4. Moderately ill - Depressive symptoms occur daily or nearly daily with substantial but bearable distress; functioning is somewhat reduced; suicidal thoughts may be present but there is usually a desire to live.
5. Markedly ill - Depressive symptoms are highly distressing; major struggle to function in important roles; active suicidal ideation may be present.
6. Severely ill - Depressive symptoms are nearly constant and highly distressing; unable to function in important life roles; active suicidal ideation may be present.
7. Among the most extremely ill patients - Depressive symptoms are continuously very severe; unable to maintain basic functioning; active suicidal thoughts usually present; hospitalization usually required.

When assessing depressive symptoms, focus on core depressive symptoms (depressed mood, anhedonia), with supporting symptoms (appetite disturbance, sleep disturbance, psychomotor agitation/retardation, fatigue, worthlessness/guilt, decreased concentration, and suicidal thoughts) as supportive of the determination/severity.

You may refer to prior provided notes of the patient, but not future notes, to help provide context.

Consider the following four sources of information, listed in no particular order:
1) The patient’s occupational, social, and daily functioning and the extent to which deficits in their functioning may be caused by depressive symptoms;
2) Patient-reported depressive symptom severity and frequency as per the HPI and mood item of mental status exam;
3) The treating clinician’s judgments and perceptions of the patient’s depressive symptom severity as expressed through mental status exam (especially the affect), the diagnosis provided (e.g. “depression in partial remission”) and narrative assessment, although some care must be taken to determine whether the mental status exam affect and listed diagnosis is truly up-to-date vs a copy-forward artifact (may compare to prior note from same patient/clinician)
4) The presence and severity of recent self-injurious behavior: Any self-injurious behavior in the past 2 weeks precludes a CGI-S lower than 3, and serious self-injurious behavior is a fast ticket to very high CGI-S scores.

When a patient could very reasonably receive either of 2 adjacent CGI-S scores, break the tie by picking the number that best represents the functional impact of their depressive symptoms, and if there is still a tie, round down.

In general, do not consider the appropriateness of the patient's symptoms for their circumstances (e.g. low mood after a recent loss) and instead rate the severity of their current depressive symptoms on an absolute scale; the only exception to this is when deciding between a 1 and a 2, remember that a patient with inappropriate depressive symptoms is not eligible for the score of 1.

We do not require depressive symptoms and their functional and behavioral consequences to be clearly attributable to MDD per se: it suffices to believe that the deficits are related to depressive symptoms which may themselves be secondary to another primary condition.

We are aiming to assess depressive severity over the course of the past 2 weeks culminating in the day of the clinical encounter. It is permissible to use information from earlier than two weeks as context when estimating their depression severity in the past 2 weeks."

*Adapted from*:

Dunlop BW, Gray J, Rapaport MH. Transdiagnostic Clinical Global Impression scoring for routine clinical settings. *Behav Sci (Basel)*. 2017;7(3):40.

### Depression-Specific Clinical Global Impression – Severity Text (*Short*)

"Clinical Global Impression - Severity (Major Depressive Disorder)

When assessing depressive symptoms, focus on core depressive symptoms (depressed mood, anhedonia), with supporting symptoms (appetite disturbance, sleep disturbance, psychomotor agitation/retardation, fatigue, worthlessness/guilt, decreased concentration, and suicidal thoughts) as supportive of the severity.

Severity Rating:
1. Normal - Depressive symptoms are rarely present and occur only in contextually appropriate circumstances. The patient reports functioning at or very close to their full capacity.
2. Borderline ill - Depressive symptoms are few in number and only intermittently present, and usually no more than mild severity. There is little or no interference in role functioning.
3. Mildly ill - Depressive symptoms are clearly present and cause distress, but there is only minimal or no reduction in functioning.
4. Moderately ill - Depressive symptoms are present every day or nearly every day but may diminish at times. Substantial distress is present but bearable. Functioning in important roles is somewhat reduced, or maintained only through high levels of perceived effort. Suicidal thoughts may be present, but there is usually a desire to live.
5. Markedly ill - Depressive symptoms are highly distressing and the patient struggles greatly to function in important life roles. Active suicidal ideation may be present.
6. Severely ill - Depressive symptoms are nearly constant and highly distressing, and the patient is unable to function in important life roles. Active suicidal ideation may be present.
7. Among the most extremely ill patients - Depressive symptoms are continuously present at a very severe level. The person is unable to maintain basic functioning. Active suicidal thoughts are usually present. Hospitalization is usually required."

*Adapted from*:

Dunlop BW, Gray J, Rapaport MH. Transdiagnostic Clinical Global Impression scoring for routine clinical settings. *Behav Sci (Basel)*. 2017;7(3):40.
